# Predicting Preclinical Cognitive Decline Using Plasma P-tau217 and *APOE* Genotype in 8,582 Individuals From Different Ethnic Groups

**DOI:** 10.64898/2026.02.06.26345774

**Authors:** Yuexuan Xu, Tamil Iniyan Gunasekaran, Yian Gu, Dolly Reyes-Dumeyer, Angel Piriz, Danurys Sanchez, Diones Rivera Mejia, Martin Medrano, Rafael A. Lantigua, Alzheimer’s Disease Neuroimaging Initiative, Lawrence Honig, Rachael Wilson, Ramiro Eduardo Rea Reyes, Jennifer J Manly, Adam M. Brickman, Corinne D. Engelman, Sterling Johnson, Sanjay Asthana, David A. Bennett, Melissa Petersen, Sid O’Bryant, Badri N. Vardarajan, Richard Mayeux

## Abstract

**Background:** Plasma phosphorylated tau217 (P-tau217), a plasma biomarker of Alzheimer’s disease (AD), can increase before overt symptoms. P-tau217 positivity is linked to the age at symptom onset in model-based predictions, not time to clinical event. Individuals with different genetic backgrounds, yet similar P-tau217 levels may differ in whether cognitive decline will occur or when it will emerge. Whether *APOE*-ε4 carrier status provides additional prognostic information beyond P-tau217 remains unclear.

**Methods:** Using data from 8,582 individuals in several multi-ethnic cohorts, we evaluated how *APOE*-ε4 carrier status modifies the risk and time to cognitive impairment associated with plasma P-tau217. Plasma P-tau217 was analyzed as a continuous measure, with positivity analyses performed secondarily. Associations of baseline P-tau217 with prevalent and incident cognitive impairment were assessed using logistic regression and Cox models, stratified by *APOE*-ε4 and in interaction models. Adjusted survival curves, restricted mean survival time, and accelerated failure time model were used to predict time to event and risk. Prognostic performance was evaluated using discrimination measures, including the AUC, incremental R², and Harrell’s C-index, and nonparametric random survival forest models.

**Findings:** Elevated P-tau217 levels were associated with subsequent cognitive impairment in both *APOE*-ε4 carriers and non-carriers, but the effects on risk and timing of cognitive impairment were significantly stronger among *APOE*-ε4 carriers. In stratified meta-analyses, increase in P-tau217 levels was associated with cognitive impairment at baseline and with incident cognitive impairment in *APOE*-ε4 carriers compared to non-carriers (OR = 2.25 vs 1.52; HR = 1.76 vs 1.26 for 1-SD increase of P-tau217 levels). Each 1-SD increase in P-tau217 levels was accompanied by a 23% shorter period to cognitive impairment among *APOE*-ε4 carriers, compared to 13% among non-carriers. Clinically relevant differences in cognitive-impairment-free survival emerged three to four years before symptom onset. Across parametric and nonparametric models, the prognostic value of P-tau217 was consistently greater among *APOE*-ε4 carriers.

**Interpretation:** Plasma P-tau217 levels and *APOE* genotypes are commercially available and can be used to estimate the years before the onset of overt cognitive impairment. These findings may also determine optimal timing for therapeutic intervention, particularly during the preclinical phase of the disease.

**Funding:** NIH

**Research in context:** *Evidence before this study:* Plasma P-tau217 has emerged as a blood biomarker of AD, and *APOE*-ε4 carrier status is the most common genetic risk factor for AD. Prior studies have shown that elevated P-tau217 levels are associated with greater risk of cognitive decline and may be elevated years before overt symptoms. However, four important questions remain unresolved. First, analytic models using P-tau217 have only estimated when cognitive impairment might occur. Second, although prior studies have examined *APOE-*ε4 and P-tau217 together in the context of amyloid pathology, it remains unclear whether *APOE-*ε4 carrier status modifies the prognosis at a given P-tau217 level, particularly for the risk and timing of cognitive impairment. Third, most prior studies have been conducted predominantly in non-Hispanic white populations, limiting the generalizability across diverse ancestry groups. Fourth, studies addressing timing have largely relied on model-based predicted age at symptom onset rather than directly observed clinically documented time-to-event.

*Added value of this study:* Using data from 8,582 individuals across several multi-ethnic cohorts, we examined whether *APOE-*ε4 carrier-status influenced the clinical prognosis using plasma P-tau217 for future cognitive impairment. By using observed clinical follow-up rather than predicted age at onset, we were able to evaluate both risk and timing in a more clinically relevant setting. Higher P-tau217 levels were associated with increased risk of cognitive impairment and earlier progression, with significantly stronger effects among *APOE-*ε4 carriers. Compared with non-carriers, *APOE-*ε4 carriers showed a higher likelihood of progression and a shorter symptom-free interval at similar P-tau217 levels. Clinically meaningful differences in impairment-free survival emerged approximately three to four years before symptom onset. Across conventional discrimination analyses and out-of-sample survival prediction, P-tau217 showed its strongest prognostic performance among *APOE-*ε4 carriers.

*Implications of all the available evidence:* Plasma P-tau217 appears to provide prognostic information not only about whether cognitive impairment is likely to occur, but also about the likely time window before symptom onset. *APOE-*ε4 carrier status refines this interpretation. Together, these findings support a more personalized prognostic framework in which the clinical meaning of a given P-tau217 level depends in part on genetic background, with potential relevance for risk stratification, trial enrichment, and timing of early intervention in diverse population groups.

## INTRODUCTION

Alzheimer’s disease (AD) is a progressive neurodegenerative disorder with a long preclinical period before the onset of overt cognitive symptoms^1^. Plasma tau phosphorylated at threonine 217 (P-tau217) is a biomarker of AD-related amyloid and tau pathology^2^ and is strongly correlated with amyloid positron emission tomography (PET) uptake^3^. Elevated plasma P-tau217 precedes clinical symptoms and predicts amyloid pathology^4,5^, and recent Alzheimer’s Association criteria^6^ have suggested including P-tau217 positivity in the definition of preclinical AD.

The apolipoprotein E (*APOE*) ε4 allele is an established genetic risk factor for AD. Individuals carrying *APOE*-ε4 develop AD pathology and symptoms earlier than non-carriers^7,8^ and progress more rapidly^9^. Because plasma P-tau217 reflects ongoing AD-related pathology, it may help identify the likely time interval before the symptom onset. The age at plasma P-tau217 positivity predicts an estimated age at onset of AD symptoms^10^ using analyses relying on model-based predicted ages rather than observed time-to-event. However, the period of time an individual remains free of cognitive impairment after biomarker assessment remains unknown. In addition, individuals with similar P-tau217 levels may differ substantially in the timing of symptoms onset according to genetic background, particularly the *APOE* ε4 status.

We hypothesize that the *APOE* genotype can further refine the prognostic value of plasma P-tau217^11^. This would provide an opportunity to identify a window for therapeutic intervention or clinical trials. Here we investigated data in 8582 individuals from seven independent multi-ethnic cohorts to determine whether *APOE* genotype improves the prognostic value of plasma P-tau217 for estimating the risk and timing of cognitive impairment.

## METHODS

An in-depth overview of all methods is provided in the Supplementary Materials. Longitudinal clinical data, *APOE* genotypes, plasma P-tau217 levels, and AD-related clinical outcomes, were included from the Wisconsin Registry for Alzheimer’s Prevention (WRAP), the Wisconsin Alzheimer’s Disease Research Center (Wisconsin-ADRC), the Alzheimer’s Disease Neuroimaging Initiative (ADNI), the Estudio Familiar de Influencia Genética en Alzheimer (EFIGA), the Washington Heights, Hamilton Heights, Inwood Columbia Aging Project (WHICAP), the Religious Orders Study/Rush Memory and Aging Project (ROSMAP), and the Health & Aging Brain Study - Health Disparities (HABS-HD) ^12–18^. The study design for each cohort has been described previously^12–19^. We harmonized WRAP and the Wisconsin-ADRC into a single Wisconsin cohort to increase sample size of symptomatic individuals. The Columbia University Medical Center Institutional Review Board approved the study. Data from each cohort was accessed with approval from the respective principal investigators and their data access committees. All analyses were conducted in accordance with the STROBE guidelines^20^.

### APOE Genotyping and Plasma P-tau217 Measurement

Genotypes were generated using high-throughput platforms across cohorts, with quality control procedures described previously^21–23^. For analysis, participants were grouped as ε4 carriers (ε3/ε4, ε4/ε4) and non-carriers (ε2/ε3, ε3/ε3). Individuals with ε2/ε2 and ε2/ε4 genotypes were excluded due to rarity and potentially protective effects of the ε2 allele.

The acquisition and processing of plasma P-tau217 for each cohort was described previously^24–29^. Validated thresholds for plasma P-tau217 positivity were not available for all cohorts thus positivity-based analyses were not used as the primary analytical approach. Instead, we used continuous P-tau217 levels as the primary exposure. For cross-sectional analyses, P-tau217 values were log₁₀-transformed and z-standardized within the full sample of each cohort to harmonize measurement scales across platforms regardless of amyloid status; for longitudinal analyses, log transformation and standardization were performed using only cognitively unimpaired individuals at baseline within each cohort. As secondary analyses, we applied a three-range positivity approach using two cohort-specific thresholds (Supplementary Table 1), one maximizing sensitivity and one maximizing specificity, to classify P-tau217 status. For cohorts without an established threshold, a proxy threshold was used.

### Clinical Endpoints Assessment

All individuals were clinically evaluated and diagnosed during study visits in each cohort^12,19,30,31^, without use of the plasma, cerebrospinal fluid, or PET biomarkers. Cohorts varied substantially in age at recruitment and number of individuals with clinically diagnosed dementia, AD, and MCI (Table 1). To increase comparability across the AD clinical spectrum and reflect the earliest changes of AD, we combined dementia, AD, and MCI as a cognitively impaired group and cognitively unimpaired individuals as normal.

**Table 1.**
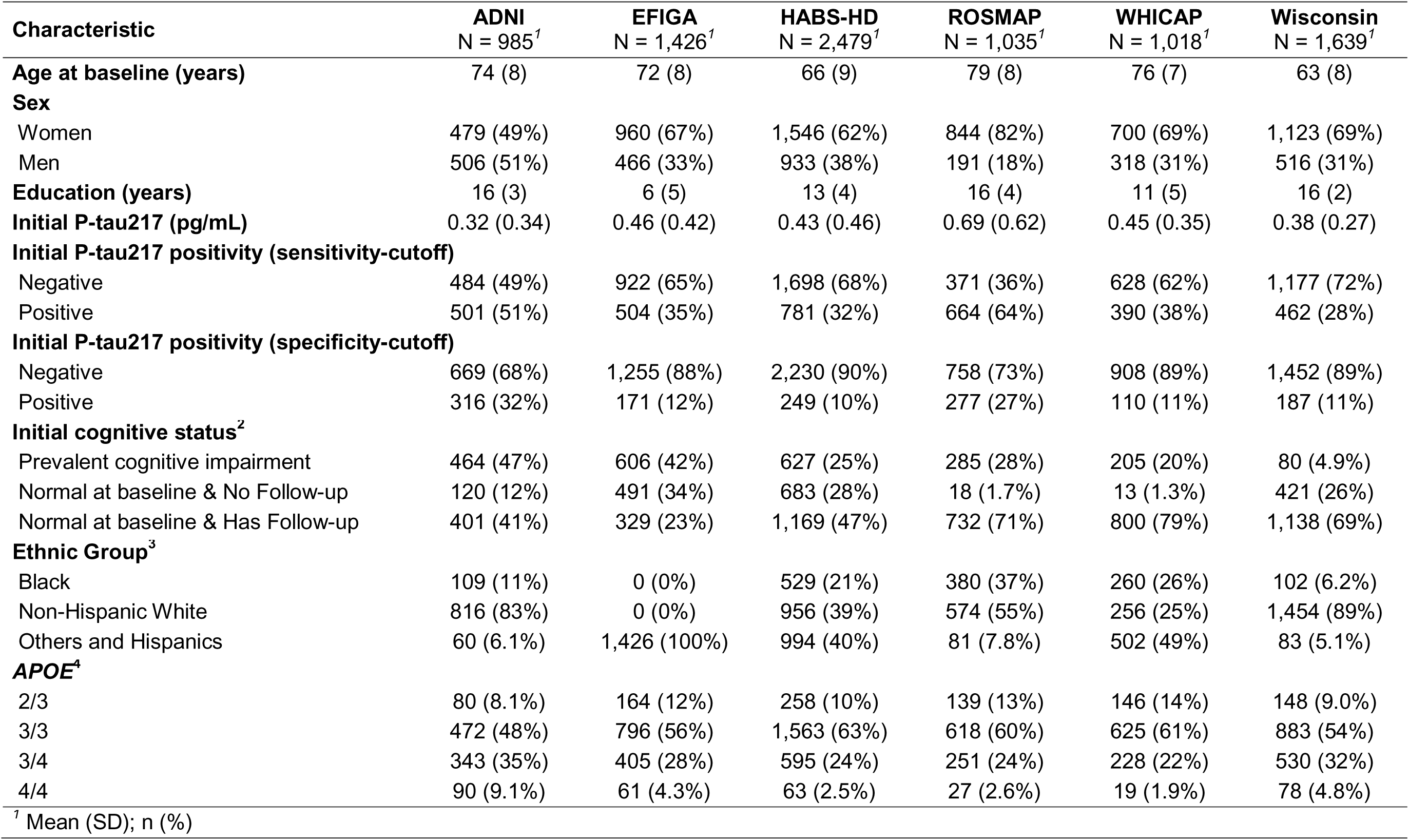
Baseline characteristics of participants included in the analysis. Table 1 presents summary statistics for the baseline characteristics of the biomarker analytic sample across cohorts. Biomarker availability and longitudinal follow-up varied by cohort; the baseline sample reflects the first available plasma biomarker measurement for each participant. For cohorts with only cross-sectional biomarker data, this represents a single time point; for longitudinal cohorts, it corresponds to the first time a biomarker was collected. ^2^ Baseline cognitive status was defined at the time when plasma biomarker data first became available for each participant. Cognitive impairment at follow-up reflects diagnoses made after this baseline assessment; any cognitive status assessments prior to biomarker availability were not considered. ^3^ Ethnic group identity was collected in detail within each cohort; however, the distribution varied substantially across studies. For example, Wisconsin and ADNI were predominantly composed of non-Hispanic White participants, followed by Black individuals, with very few Hispanic or other ethnic groups. In contrast, EFIGA included only Hispanic participants, while WHICAP had a more balanced ethnic composition. To facilitate analysis and ensure adequate sample sizes within each category, we combined Hispanic and other ethnic groups into a single category. This grouping also helped stabilize regression models by reducing sparse categories and improving model convergence. ^4^ *APOE* ε2/ε2 and ε2/ε4 genotypes were excluded due to their low frequency and the potentially protective effects of the ε2 allele, which may confound interpretation of *APOE*-ε4–related risk associations.

| Characteristic | ADNI<br>N = 985<sup>†</sup> | EFIGA<br>N = 1,426<sup>†</sup> | HABS-HD<br>N = 2,479<sup>†</sup> | ROSMAP<br>N = 1,035<sup>†</sup> | WHICAP<br>N = 1,018<sup>†</sup> | Wisconsin<br>N = 1,639<sup>†</sup> |
| --- | --- | --- | --- | --- | --- | --- |
| **Age at baseline (years)** | 74 (8) | 72 (8) | 66 (9) | 79 (8) | 76 (7) | 63 (8) |
| **Sex** |  |  |  |  |  |  |
| Women | 479 (49%) | 960 (67%) | 1,546 (62%) | 844 (82%) | 700 (69%) | 1,123 (69%) |
| Men | 506 (51%) | 466 (33%) | 933 (38%) | 191 (18%) | 318 (31%) | 516 (31%) |
| **Education (years)** | 16 (3) | 6 (5) | 13 (4) | 16 (4) | 11 (5) | 16 (2) |
| **Initial P-tau217 (pg/mL)** | 0.32 (0.34) | 0.46 (0.42) | 0.43 (0.46) | 0.69 (0.62) | 0.45 (0.35) | 0.38 (0.27) |
| **Initial P-tau217 positivity (sensitivity-cutoff)** |  |  |  |  |  |  |
| Negative | 484 (49%) | 922 (65%) | 1,698 (68%) | 371 (36%) | 628 (62%) | 1,177 (72%) |
| Positive | 501 (51%) | 504 (35%) | 781 (32%) | 664 (64%) | 390 (38%) | 462 (28%) |
| **Initial P-tau217 positivity (specificity-cutoff)** |  |  |  |  |  |  |
| Negative | 669 (68%) | 1,255 (88%) | 2,230 (90%) | 758 (73%) | 908 (89%) | 1,452 (89%) |
| Positive | 316 (32%) | 171 (12%) | 249 (10%) | 277 (27%) | 110 (11%) | 187 (11%) |
| **Initial cognitive status<sup>2</sup>** |  |  |  |  |  |  |
| Prevalent cognitive impairment | 464 (47%) | 606 (42%) | 627 (25%) | 285 (28%) | 205 (20%) | 80 (4.9%) |
| Normal at baseline & No Follow-up | 120 (12%) | 491 (34%) | 683 (28%) | 18 (1.7%) | 13 (1.3%) | 421 (26%) |
| Normal at baseline & Has Follow-up | 401 (41%) | 329 (23%) | 1,169 (47%) | 732 (71%) | 800 (79%) | 1,138 (69%) |
| **Ethnic Group<sup>3</sup>** |  |  |  |  |  |  |
| Black | 109 (11%) | 0 (0%) | 529 (21%) | 380 (37%) | 260 (26%) | 102 (6.2%) |
| Non-Hispanic White | 816 (83%) | 0 (0%) | 956 (39%) | 574 (55%) | 256 (25%) | 1,454 (89%) |
| Others and Hispanics | 60 (6.1%) | 1,426 (100%) | 994 (40%) | 81 (7.8%) | 502 (49%) | 83 (5.1%) |
| **APOE<sup>4</sup>** |  |  |  |  |  |  |
| 2/3 | 80 (8.1%) | 164 (12%) | 258 (10%) | 139 (13%) | 146 (14%) | 148 (9.0%) |
| 3/3 | 472 (48%) | 796 (56%) | 1,563 (63%) | 618 (60%) | 625 (61%) | 883 (54%) |
| 3/4 | 343 (35%) | 405 (28%) | 595 (24%) | 251 (24%) | 228 (22%) | 530 (32%) |
| 4/4 | 90 (9.1%) | 61 (4.3%) | 63 (2.5%) | 27 (2.6%) | 19 (1.9%) | 78 (4.8%) |

<sup>†</sup> Mean (SD); n (%)

### Statistical Analysis

WHICAP and EFIGA had only cross-sectional plasma P-tau217 biomarker data, whereas ADNI, ROSMAP, and HABS-HD had limited follow-up, and Wisconsin had more extensive follow-up. All cohorts, however, included longitudinal clinical assessment data. To maximize comparability across cohorts, baseline was defined as the first visit with plasma biomarker measurement.

We characterized baseline plasma P-tau217 levels and examined their cross-sectional association with cognitive impairment. We subsequently summarized P-tau217 levels by cognitive status, age group (<=70 and >70), and *APOE*-ε4 status. Cohort-specific associations between baseline P-tau217 and cognitive impairment were evaluated using logistic regression adjusted for age, sex, education, ethnic group, and *APOE* genotype. To assess how associations differed by *APOE*-ε4 status, analyses were repeated within *APOE*-ε4 strata and the *APOE*-ε4*P-tau217 interaction was formally tested.

We next examined the longitudinal prognostic value of baseline P-tau217 for incident cognitive impairment among individuals who were cognitively unimpaired at baseline. Cox proportional hazards models adjusted for age, sex, education, ethnic group, and *APOE* genotype were used to assess the association between baseline P-tau217 levels and subsequent onset of cognitive impairment. Incident events were dated from baseline to onset of cognitive impairment; all others were censored at last evaluation. The proportional hazards assumption was assessed using Schoenfeld residuals^32^. We repeated the longitudinal analyses stratified by *APOE*-ε4 status and tested the *APOE*-ε4 *P-tau217 interaction to determine whether *APOE*-ε4 modified the prognostic value of baseline P-tau217. Random-effects meta-analyses were used to combine cohort-specific estimates for the cross-sectional association between baseline P-tau217 and cognitive impairment, and for the hazard ratio (HR) relating baseline P-tau217 to incident cognitive impairment, overall and within *APOE*-ε4 strata.

To characterize timing of progression and prognosis, we conducted pooled post hoc analyses across all cohorts, including cohort as a covariate to account for between-cohort differences. Parallel post hoc analyses using P-tau217 positivity defined by cohort-specific thresholds were performed as secondary analyses and are reported in Supplement. Covariate-adjusted survival curves were used to identify the earliest divergence in cognitive-impairment free survival across clinically relevant P-tau217 levels (e.g., the mean and +1 standard deviation [SD] within each cohort)^33–39^. To quantify differences in time to impairment, we then compared the time at which adjusted survival reached 0.75 (S(t) = 0.75), a measure less influenced by incomplete follow-up in younger cohorts that did not reach the median survival time (e.g., Wisconsin). The proportional hazards assumption was violated in the pooled continuous P-tau217 models, likely due to between-dataset heterogeneity, thus these pooled Cox models were stratified^24^.

We quantified average time free of cognitive impairment over time using restricted mean survival time (RMST) and fitted accelerated failure time (AFT) models to formally evaluate whether baseline P-tau217, and its interaction with *APOE*-ε4, were associated with earlier onset of cognitive impairment. AFT models do not require the proportional hazards assumption, but they provided time-ratio estimates that complemented the Cox-based analyses. For the survival curve and RMST analyses, we applied the Benjamini–Yekutieli false discovery rate (BY-FDR) procedure, which accommodates dependence among tests. For comparability across strata, survival and RMST estimates were truncated at 14 years because approximately 99% of observations occurred within this period.

Finally, we evaluated whether combining P-tau217 with *APOE*-ε4 improved prognostic performance using area under the ROC curve (AUC) and incremental R² in the cross-sectional models, and Harrell’s concordance index (C-index) in the longitudinal analyses. In the pooled longitudinal sample, stratification was applied for covariates that violated the proportional hazards assumption.

As sensitivity analyses, we repeated the survival, RMST, and discrimination analyses within each cohort. We also explored predictive performance over follow-up using non-parametric random forest models^40^. As an exploratory post-hoc analysis, we examined *APOE* associations with P-tau217 across different disease stages and with disease status, jointly defined by clinical cognitive impairment and P-tau217 positivity, to facilitate interpretation of between-cohort heterogeneity. All analyses were performed in R (v4.2.2).

## RESULTS

### Plasma P-tau217 and Cognitive Impairment at Baseline

Descriptive statistics at baseline are presented in Table 1, and those for the subset in the longitudinal analysis are presented in Supplementary Table 2. Overall, P-tau217 concentrations were higher in cognitively impaired individuals compared to individuals without cognitive impairment (Figure 1). Differences were more pronounced among *APOE*-ε4 carriers, including younger participants, with greater elevations in both magnitude and statistical significance across most datasets.

**Figure 1.**
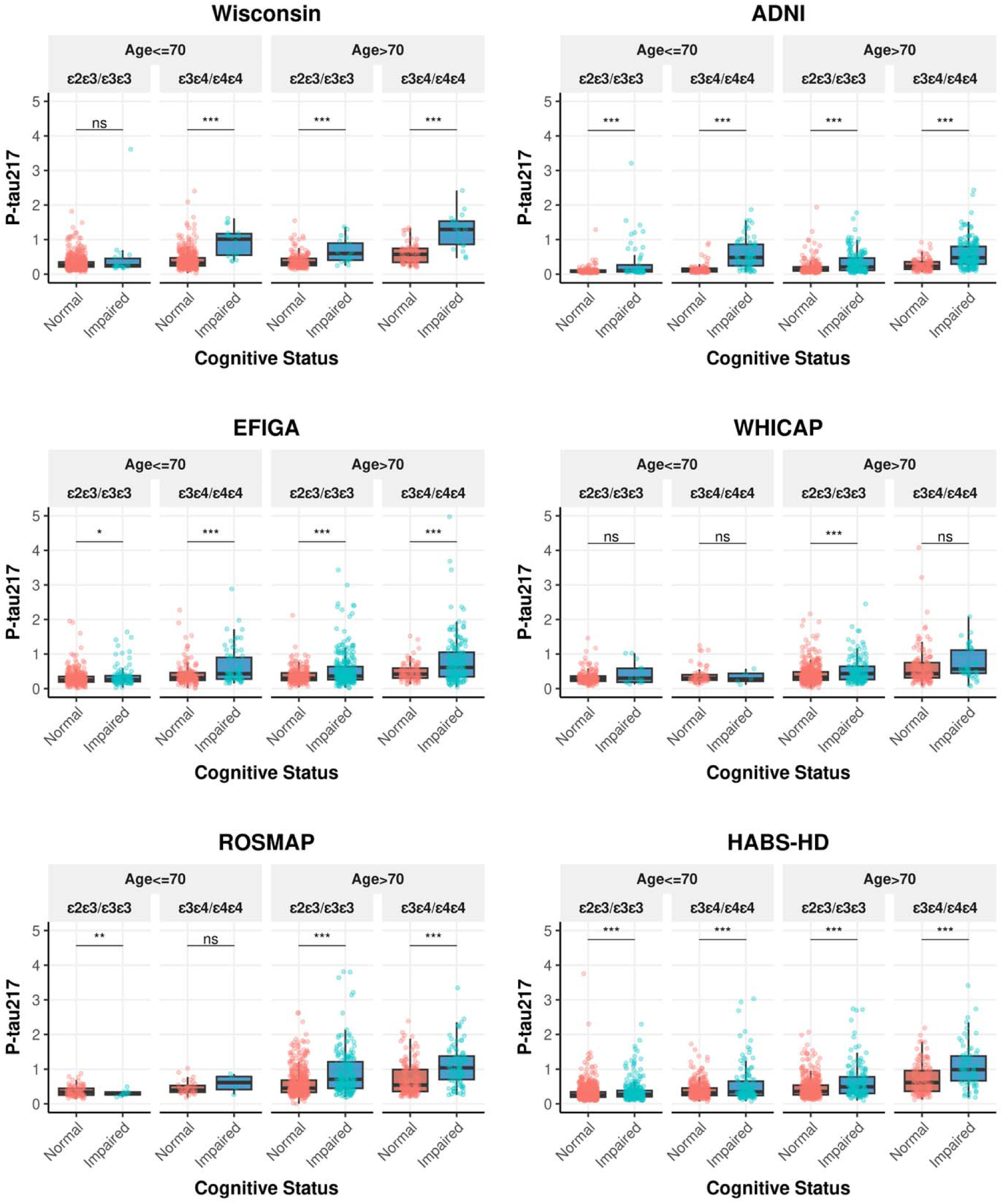
Raw plasma P-tau217 levels by cognitive status, *APOE* genotype, and age group across cohorts. Figure 1 presents plasma P-tau217 levels by cognitive status, *APOE* genotype, and age group across cohorts. Analyses were conducted at the time of the first available biomarker measurement. Data are shown separately for the Wisconsin, ADNI, EFIGA, WHICAP, ROSMAP, and HABS-HD cohorts. Cognitive status was classified as Normal (cognitively unimpaired) or Impaired (mild cognitive impairment [MCI] or Alzheimer’s disease [AD]). Participants were further stratified by *APOE* genotype (ε2/ε3, ε3/ε3 or ε3/ε4, ε4/ε4) and by age group (<=70 years vs >70 years). Boxplots display the median, interquartile range, and individual participant values (dots). Differences in plasma P-tau217 levels between groups were assessed using Welch’s t-test, with significance levels denoted as ns (not significant), * p < 0.1, ** p < 0.05, and *** p < 0.01.

Statistically significant associations between P-tau217 and baseline cognitive impairment were observed in the Wisconsin, ADNI, EFIGA, ROSMAP, and HABS-HD datasets (Figure 2A). In meta-analyses across cohorts, each 1-unit increase in within-cohort standardized, log-transformed P-tau217 level was associated with higher odds of cognitive impairment (OR = 1.77, 95% CI: 1.42-2.20). When stratified by *APOE*-ε4 status, the association remained significant in both groups but was stronger among *APOE*-ε4 carriers (OR = 2.25, 95% CI: 1.52-3.34) than non-carriers (OR = 1.52, 95% CI: 1.35-1.72). Consistent with this pattern, the combined-sample model showed a significant P-tau217 by *APOE*-ε4 interaction (*p* < 0.001). Similar interaction effects were observed in Wisconsin and ADNI and were marginal in EFIGA (Figure 2B–2C).

**Figure 2.**
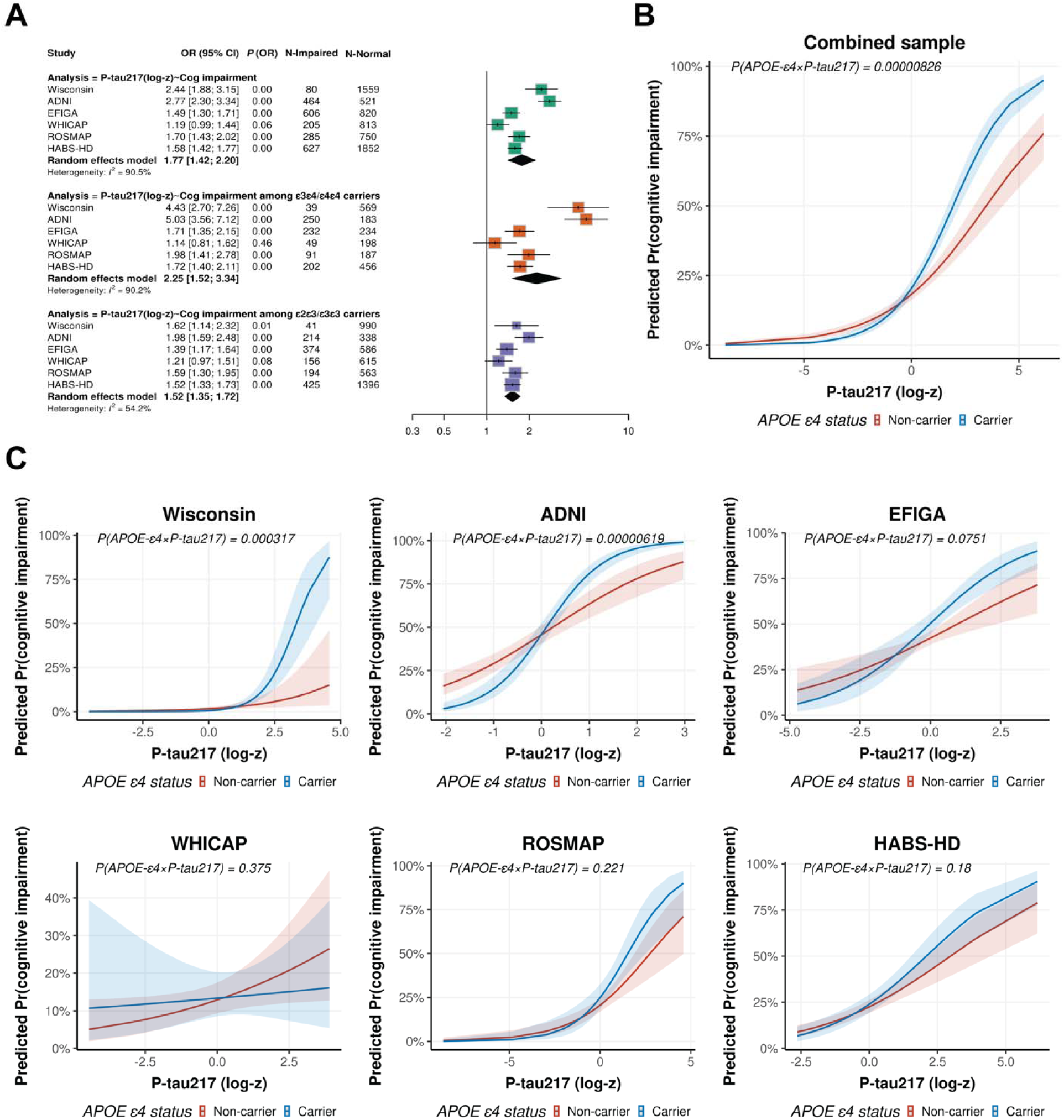
Cohort-specific associations between plasma P-tau217 and clinical cognitive impairment, with meta-analysis estimates in the full sample and stratified subgroups (A), and *APOE*-ε4 × P-tau217 interactions in the combined sample (B) and individual cohorts (C). Figure 2 presents cohort-specific associations between harmonized plasma P-tau217 and clinical cognitive impairment, along with meta-analytic estimates in the full sample and stratified subgroups (A). We also evaluated *APOE*-ε4 × P-tau217 interactions in the combined sample, which included all cohorts (B), as well as within each individual cohort (C). Analyses were conducted at the time of first available biomarker measurement. For cohorts with only cross-sectional data, this corresponds to a single time point; for longitudinal cohorts, it reflects the earliest available measure. Plasma P-tau217 was harmonized using log₁₀ transformation and z-standardization within each cohort. Associations were tested using logistic regression models adjusted for age, sex, education, ethnic group, cohort (in the combined sample only), and *APOE* genotype; Within each full or stratified subgroup, random-effects meta-analyses were performed using cohort-specific ORs. Cognitive impairment was defined as a diagnosis of MCI or AD.

### Baseline P-tau217 and Longitudinal Risk of Cognitive Impairment

Higher baseline P-tau217 levels were associated with an increased risk of incident cognitive impairment in the Wisconsin, ADNI, WHICAP, ROSMAP, and HABS-HD datasets (Figure 3A). In meta-analyses across cohorts, each 1-unit increase in within-cohort standardized, log-transformed P-tau217 level was associated with incident cognitive impairment (HR = 1.41, 95% CI: 1.22-1.64). When stratified by *APOE*-ε4 status, the association was stronger among *APOE*-ε4 carriers (HR = 1.76, 95% CI: 1.36-2.26) than among non-carriers (HR = 1.26, 95% CI: 1.12-1.42). Consistent with this pattern, the pooled sample showed a statistically significant P-tau217 by *APOE*-ε4 interaction (*p* < 0.001). In cohort-specific analyses, significant interactions were observed only in Wisconsin and ADNI, both of which were enriched for non-Hispanic White participants (Supplementary Table 3). Results from positivity-based analyses are presented in Supplementary Figure 1.

**Figure 3.**
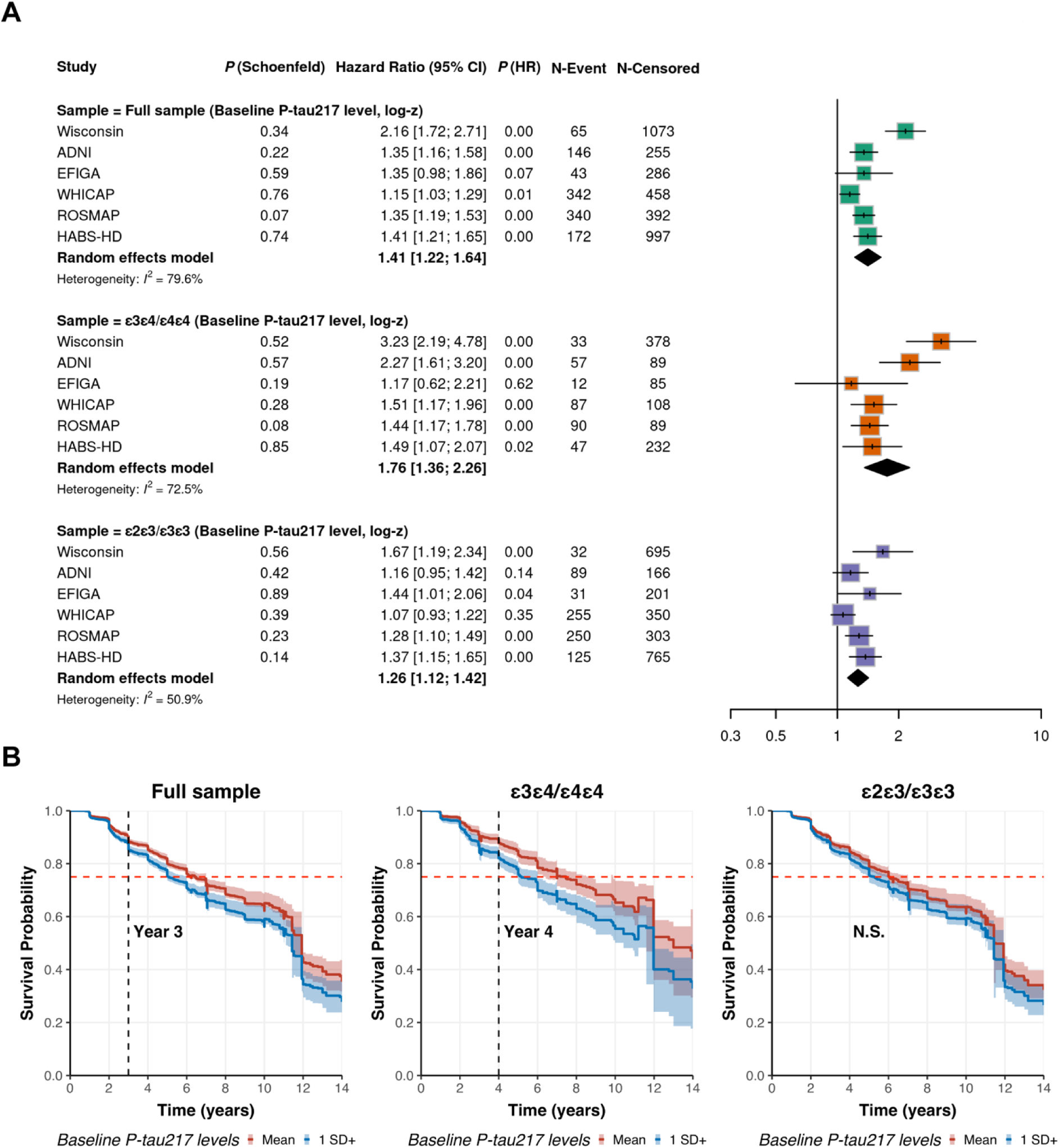
Cohort-specific and meta-analytic hazard ratios for incident cognitive impairment in the full sample and *APOE*-stratified subgroups using harmonized continuous P-tau217 measures, along with adjusted survival probabilities illustrating the temporal dynamics of P-tau217 in the pooled cohort. Figure 3 presents cohort-specific hazard ratios for incident cognitive impairment in the full sample and *APOE*-stratified subgroups using harmonized continuous P-tau217 measures, along with meta-analytic estimates (Figure 3A) and adjusted survival probabilities (Figure 3B) illustrating the temporal dynamics of P-tau217 in the pooled cohort. Given variability in longitudinal biomarker availability across cohorts, a standardized longitudinal analysis framework was applied. For each participant, baseline was defined as the time of first available biomarker measurement. In cohorts with only cross-sectional biomarker data, this represents a single time point, whereas in longitudinal cohorts it corresponded to the earliest biomarker collection. Clinical follow-up data were linked to each participant’s biomarker baseline, and only visits occurring after this time point were included. Analyses were restricted to individuals who were cognitively unimpaired at baseline and had follow-up data. Incident cognitive impairment was defined as conversion from cognitively unimpaired to either MCI or AD. Time-to-event was defined as the interval from baseline to the date of diagnosis for converters or to the last available follow-up for censored cases. Cox proportional hazards models were used, adjusting for age, sex, education, ethnicity, and *APOE* genotype. The proportional hazards assumption was tested using Schoenfeld residuals. Within each full or stratified subgroup, random-effects meta-analyses were conducted using cohort-specific estimates. Continuous P-tau217 was harmonized using log₁₀ transformation and z-standardization among cognitively unimpaired individuals at baseline within each cohort. P(Schoenfeld) indicates the p-value from the Schoenfeld global test for proportional hazards. For the adjusted survival curve and to maximize statistical power, data from all cohorts were pooled. Log-transformation and standardization of continuous P-tau217 levels were performed within each individual cohort and then combined without further standardization; thus, the values in the combined cohort reflect cohort-specific standardized units rather than a single unified global scale. We addressed violations of the proportional hazards’ assumption in the pooled sample by stratifying on the covariates that failed the Schoenfeld global test before plotting the survival curves. We then plotted covariate-adjusted survival curves at pre-defined P-tau217 levels (e.g., the cohort-specific mean and +1 SD) to allow clinically meaningful comparisons, as +1 SD reflects a clearly elevated P-tau217 level relative to the cohort average. The black vertical dashed line marks the earliest follow-up year at which the two adjusted survival curves began to diverge. The red dashed line indicates the follow-up time at which adjusted cognitive-impairment-free survival reached 0.75.

### Interval between High P-tau217 levels and Cognitive Impairment

In pooled analyses using continuous P-tau217, adjusted survival curves showed little difference in cognitive-impairment-free survival during the first 3 years between individuals with P-tau217 levels 1 SD above the mean and those at the mean, suggesting a delay between elevated baseline P-tau217 and clinically detectable cognitive impairment (BY-FDR–adjusted, Figure 3B). However, at year 4, *APOE*-ε4 carriers with P-tau217 1 SD above the mean began to diverge from those at the mean, indicating the onset of cognitive impairment, whereas no clear separation was observed among non-carriers. RMST analyses showed a similar overall pattern (Supplementary Figure 2), supporting a delayed emergence of the association between higher P-tau217 and cognitive-impairment-free survival, particularly among *APOE*-ε4 carriers.

Higher P-tau217 levels were associated with earlier cognitive impairment, with a larger shift in timing among *APOE*-ε4 carriers than among non-carriers. Relative to individuals at the mean P-tau217 level, those with P-tau217 levels 1 SD above the mean reached the 75% cognitive-impairment–free survival threshold 1.8 years earlier among *APOE*-ε4 carriers and 0.8 years earlier among non-carriers. Consistent with these findings, pooled AFT models showed a significant *APOE*-ε4 by P-tau217 interaction, indicating that the association between higher P-tau217 and earlier cognitive impairment was stronger among *APOE*-ε4 carriers than among non-carriers (Supplementary Table 4). Specifically, each 1-SD increase in within-cohort standardized, log-transformed P-tau217 level was associated with an 23% shorter time to cognitive impairment among *APOE*-ε4 carriers, compared with an 13% shorter time among non-carriers. Similar patterns were observed in the positivity analyses (Supplementary Figures 1-2, Supplementary Table 4), and results were broadly consistent across cohorts (Survival curve: Supplementary Figures 3–5; RMST: Supplementary Figures 6-8).

### Predictive Discrimination of P-tau217 by APOE-ε4 Status

Discrimination analyses showed strong baseline predictive performance for continuous P-tau217 (AUC = 0.795) and *APOE*-ε4 (AUC = 0.774). Notably, continuous P-tau217 among *APOE*-ε4 carriers yielded the highest discrimination (AUC = 0.835), exceeding that of the additive model including both continuous P-tau217 and *APOE*-ε4, suggesting that stratification by *APOE*-ε4 status may better capture risk differences than modeling the two markers additively. Similar patterns were observed for incremental R² and longitudinal C-index estimates (Figure 4). These findings were generally consistent across most cohorts. Results from the positivity-based analyses are presented in Supplementary Figure 9, and cohort-specific results are shown in Supplementary Figures 10-15. Findings were also generally consistent when positivity was defined using a uniform cut-off derived from the Wisconsin cohort and applied across datasets (Supplementary Figures 16-17). In a separate leave-one-cohort-out external validation analysis, random survival forest models including P-tau217 and covariates showed the best predictive performance among *APOE*-ε4 carriers compared with the other models. Time-dependent AUC values exceeded 0.75 at most time points, and the magnitude of performance improvement increased with longer follow-up time across cohorts (Supplementary Figure 18).

**Figure 4.**
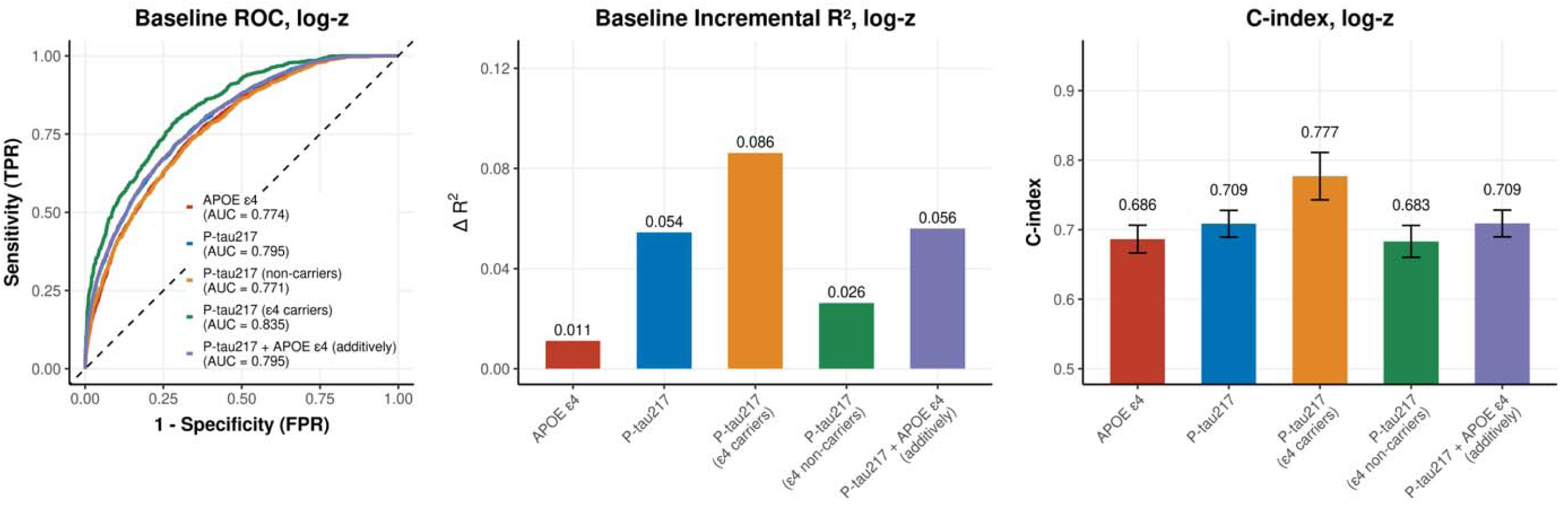
Baseline and longitudinal prognostic performance of *APOE*-ε4 carrier status and plasma P-tau217 levels, including P-tau217 stratified by *APOE*-ε4 carrier status, for cognitive impairment in the pooled cohort, as assessed by AUC, incremental R², and Harrell’s C-index. Figure 4 shows baseline and longitudinal prognostic performance of plasma P-tau217 levels for cognitive impairment in the pooled cohort. Receiver operating characteristic (ROC) curves were obtained from logistic regression models adjusted for age, sex, education, cohort, and ethnic group. Models included *APOE*-ε4 status with covariates; plasma P-tau217 with covariates; plasma P-tau217 and *APOE*-ε4 entered additively with covariates; and the plasma P-tau217 with covariates fitted separately within *APOE*-ε4 carrier and non-carrier groups. Incremental Nagelkerke’s R² was defined as the increase in model fit when adding the predictor or the predictors of interest to the covariate-only model. Harrell’s C-indices were obtained from Cox proportional hazards models adjusted for the same covariates and specified analogously.

We examined cross-cohort and cross-ancestry heterogeneity. Study-specific analyses showed that *APOE*-ε4 was significantly associated with P-tau217 levels in all cohorts, including among cognitively unimpaired individuals within each cohort (Supplementary Figure 19A). *APOE*-ε4 was also associated with P-tau217 levels among biomarker negative individuals, regardless of whether negativity was defined using sensitivity- or specificity-based threshold. Similarly, *APOE*-ε4 was associated with P-tau217 positivity under both threshold definitions (Supplementary Figure 19B). Notably, these associations showed essentially no heterogeneity across cohorts among cognitively unimpaired and P-tau217–negative individuals when the sensitivity-based cutoff was applied.

The proportion of cognitively impaired who were P-tau217 positive differed substantially across ancestries. Under the sensitivity-based cutoff, 74% of cognitively impaired individuals among non-Hispanic white participants were P-tau217 positive, compared with 52% among Black individuals, and 45% among Hispanic individuals. Under the specificity-based cutoff, the corresponding proportions were 51%, 24%, and 22%, respectively. When *APOE-*ε4 associations were examined across groups jointly defined by clinical status and P-tau217 positivity, the strongest associations were consistently observed in P-tau217-positive groups across ancestries. In contrast, the association between *APOE-*ε4 and clinically impaired but P-tau217-negative individuals was substantially weaker, with only minor variation by ancestry (Supplementary Table 5).

## DISCUSSION

In this study of 8,582 individuals from seven cohorts with diverse ethnic backgrounds, we found that plasma P-tau217 informs, not only the risk of cognitive impairment, but provides time window over which cognitive impairment may emerge that differs by *APOE* genotype. Although higher baseline P-tau217 was associated with prevalent cognitive impairment, greater risk of incident impairment, and earlier onset of symptoms overall, individuals with similar P-tau217 levels did not follow the same clinical trajectory. In particular, the prognostic effect of elevated P-tau217 was stronger among *APOE-*ε4 carriers than non-carriers, indicating that *APOE* genotype augments the clinical interpretation of a given P-tau217 level. Importantly, elevated baseline P-tau217 preceded clinically detectable divergence in cognitive impairment free survival time by approximately three years in the full sample and about four years among *APOE-*ε4 carriers. Together, these findings suggest that P-tau217 alone does not fully capture near-term progression risk, and that incorporating *APOE* genotype improves estimations of risk and timing of cognitive impairment. These conclusions were further supported by complementary prediction analyses, in which plasma P-tau217 showed the strongest discrimination among *APOE-*ε4 carriers, outperforming either marker alone, their additive combination, and corresponding models among non-carriers, in baseline logistic AUC analyses and in out-of-sample random survival forest prediction of future cognitive impairment.

Elevated plasma P-tau217 is widely viewed as a marker of underlying AD pathology, but our findings suggest that this prognostic interpretation is not uniform across individuals. At a similar level of baseline P-tau217, *APOE-*ε4 carriers have a steeper transition toward cognitive impairment, a shorter impairment-free interval, and a higher risk of progression than non-carriers. These findings indicate that comparable P-tau217 levels may reflect different stages or rates of disease progression depending on genetic background. One possible interpretation is *APOE-*ε4 may accelerate downstream consequences of AD pathology. Prior studies have suggested that *APOE-*ε4 impairs lipid transport in the brain, promotes glial lipid droplet accumulation, reduces microglial phagocytic function, and increases neurotoxic responses^41–43^. All these mechanisms could favor amyloid buildup, tau spread, and subsequent neuronal injury. Our findings are consistent with previous studies showing faster progression and greater risk associated with elevated P-tau217 among *APOE-*ε4 carriers^11^.

The observations here show elevated P-tau217 preceding clinically detectable separation particularly among *APOE-*ε4 carriers. This indicates that plasma P-tau217 can be used to identify a clinically meaningful pre-symptomatic window albeit when AD pathology accumulates. In contrast, there was no clear separation was observed among non*-APOE-*ε4 carriers over the same interval. Prior studies have linked plasma P-tau217 positivity to age at symptoms onset, supporting the idea that plasma P-tau217 carries temporal information about disease progression^10^. However, these studies were mainly prediction-focused and asked when symptoms may occur. Our results add a more clinically relevant perspective by showing the earliest time point at which elevated baseline P-tau217 becomes associated with measurable divergence in cognitive outcomes. In this way, the lag window is defined not by predicted symptom-onset age, but by the emergence of detectable differences in cognitive impairment during follow-up. This provides a conservative estimate of the preclinical interval before symptoms become clinically apparent, which would be useful for prevention trials and early intervention strategies using existing AD biomarkers.

The prognostic value of plasma P-tau217 for cognitive impairment varied across cohorts with different ethnic compositions, appearing more robust in non-Hispanic white individuals compared to ethnically diverse individuals, consistent with previous findings^44^. This heterogeneity could partly reflect differences in the pathological basis of cognitive impairment across these cohorts. Cognitive impairment is a syndromic outcome rather than a pathology-specific diagnosis, and the extent to which it reflects underlying AD pathology may differ by cohort and ancestral composition. In cohorts where clinically defined impairment more closely reflects underlying AD, elevated P-tau217 may provide a clearer estimate of the interval before clinically detectable divergence in outcomes, whereas in cohorts with greater pathological heterogeneity, the corresponding lag period may be less distinct.

A related pattern was observed for the modifying effect of *APOE-*ε4 on the prognostic value of P-tau217. Although the strength of this modification varied across cohorts and ancestral groups, *APOE-*ε4 associations with P-tau217 was consistent at earlier disease stages, before cognitive symptoms had occurred, and became more heterogeneous once symptomatic individuals were included. This pattern indicates that cross-cohort differences are less likely to arise from the relationship between *APOE* and early AD-related biomarker changes themselves, and more likely to emerge at the stage of clinically defined impairment. This may also help explain the previously observed heterogeneity in *APOE* associations with clinically defined AD when biomarker information was not incorporated into diagnosis^45^. Consistent with this interpretation, *APOE* associations were stronger among P-tau217-positive individuals regardless of clinical diagnosis but were weaker among clinically impaired individuals who were P-tau217-negative. The *APOE* association in P-tau217-positive impaired individuals, relative to cognitively unimpaired biomarker negative controls, was similar to that reported in neuropathologically confirmed AD, indicating that this biomarker-defined group more closely reflects underlying AD pathology^46^.

Our study has some limitations. First, we measured P-tau217 at a single time point in most cohorts, which prevented us from capturing longitudinal within-person changes and our ability to define the lag period in relation to biomarker progression itself. Second, P-tau217 positivity may have clinical utility because binary classification could provide a more intuitive framework for risk stratification than continuous biomarker analyses alone. However, threshold for positivity likely varied across cohorts because of assay and population differences, and appropriate cutoffs remain uncertain in several settings. As a result, binary classification may have introduced additional heterogeneity and reduced the robustness and generalizability of cutoff-based estimates across cohorts. We therefore treated positivity analyses as secondary, while recognizing that broadly validated thresholds would be important for future clinical implementation. Third, adjusted survival curves and RMST analyses suggested that the effect of P-tau217 on risk of cognitive impairment may not be constant over time. Limited sample sizes and events within individual cohorts, together with substantial heterogeneity in baseline hazards and follow-up structures, made precise estimation of time-dependent hazard ratios challenging, even in the combined sample. Nevertheless, the main findings were robust in accelerated failure time models, which provided a complementary framework for assessing differences in timing of progression without relying on the proportional hazards assumption. Fourth, the present study focused specifically on the modifying effect of *APOE*-ε4 on the prognostic value of P-tau217. However, *APOE*-ε4 represents only a single genetic factor, and future studies should examine whether broader genetic profiles, including polygenic risk scores and other susceptibility variants, to further refine the relationship between P-tau217 and the risk and timing of cognitive impairment.

Assays for plasma P-tau217 and *APOE* genotyping are widely commercially available. As demonstrated, when used together, plasma P-tau217 and *APOE* genotyping provide robust early predictions of cognitive impairment, particularly among *APOE*-ε4 carriers. This “not if, but when” observation makes it possible to identify at-risk individuals years before symptom onset, thereby defining a potential window for therapeutic intervention. Although prognostic strength for clinical diagnosis varied by ethnicity, the relationship between P-tau217 and the underlying biological process appeared consistent across ethnic groups, supporting its broad clinical utility for early detection and treatment planning.

## Data availability statement

The data analyzed in this study are subject to the following licenses/restrictions: datasets can be requested through formal research applications to the Washington Heights-Inwood Columbia Aging Project and the Genetic Studies of Alzheimer’s Disease in Caribbean Hispanics (EFIGA). Requests to access these datasets should be directed to the Columbia University Alzheimer’s Disease Research Center: https://www.neurology.columbia.edu/research/research-centers-and-programs/alzheimers-disease-research-center-adrc/investigators/investigator-resources.

Data may also be requested from the Wisconsin Registry for Alzheimer’s Prevention and the Wisconsin Alzheimer’s Disease Research Center via: https://wrap.wisc.edu/data-requests-2/, from the Alzheimer’s Disease Neuroimaging Initiative (ADNI): https://adni.loni.usc.edu/data-samples/adni-data/, from the Health & Aging Brain Study (HABS-HD): https://apps.unthsc.edu/itr/our/, and from the Religious Orders Study and Memory and Aging Project (ROSMAP): https://www.radc.rush.edu/requests.htm.

## Ethics statement

The studies involving humans were approved by the Institutional Review Board of the Columbia University Medical Center. The studies were conducted in accordance with the local legislation and institutional requirements. Subjects from WRAP, Wisconsin-ADRC, WHICAP, EFIGA, ROSMAP, and HABS-HD have provided signed informed consent before participation.

## Author contributions

YX, BV, and RM contributed to the study design. YX, TG, RM, BV, DR, LH, and YG prepared the data and performed the data analysis. YX, BV, and RM drafted the manuscript. All authors critically reviewed the manuscript and have approved the final version.

## Acknowledgements

We acknowledge the participants from the Washington Heights-Inwood Columbia Aging Project (WHICAP), the Genetic Studies of Alzheimer’s Disease in Caribbean Hispanics (EFIGA), the Wisconsin Registry for Alzheimer’s Prevention (WRAP), the Wisconsin Alzheimer’s Disease Research Center (Wisconsin ADRC), the Health & Aging Brain Study – Health Disparities (HABS-HD), the Religious Orders Study and Rush Memory and Aging Project (ROSMAP), and the Alzheimer’s Disease Neuroimaging Initiative (ADNI) for their invaluable contributions and dedication. This study would not be possible without their continued participation. We also thank the researchers and study teams involved in WHICAP, EFIGA, WRAP, the Wisconsin ADRC, HABS-HD, ROSMAP, and ADNI for their longstanding efforts in data collection and subject follow-up over the years. The authors wish to acknowledge and thank the physicians in the Dominican Republic, Belisa Soriano, MD, Yahaira Franco, MD, Zoraida Dominguez Coronado, MD, Patricia Recio, MD for their help recruiting individuals in the EFIGA study. Author YX would also like to thank Yuanyuan Qiao for her patience, support, and assistance with proofreading this study.

