## Supplementary Files for "Predicting Preclinical Cognitive Decline Using Plasma P-tau217 and *APOE* Genotype in 8,582 Individuals From Different Ethnic Groups"

*Data used in the preparation of this article were obtained from the Alzheimer’s Disease Neuroimaging Initiative (ADNI) database (adni.loni.usc.edu). The investigators within ADNI contributed to the design and implementation of ADNI and/or provided data but did not participate in the analysis or writing of this report. A complete listing of ADNI investigators can be found at: <https://adni.loni.usc.edu/wp-content/uploads/how_to_apply/ADNI_Acknowledgement_List.pdf>

^**^**Correspondence:**

Richard Mayeux, MD

Neurological Institute

Columbia University

710 West 168^th^ Street

New York, NY 10032

**Supplementary Methods**

*Cohort description*

Wisconsin Registry for Alzheimer’s Prevention: The Wisconsin Registry for Alzheimer’s Prevention (WRAP) is an ongoing prospective cohort study that follows more than 1,500 cognitively healthy, English-speaking adults who were 40–65 years old at enrollment^1^. The WRAP cohort was intentionally enriched for individuals with a parental history of Alzheimer’s disease (AD) to increase the likelihood of observing late-onset AD pathology and cognitive decline over time. More than 70% of participants had a parent with autopsy-confirmed or probable AD, as defined by the NINCDS-ADRDA criteria. Recruitment began in 2001, with the first follow-up assessment conducted four years later and subsequent visits taking place every two years.

Wisconsin Alzheimer’s Disease Research Center: Established in 2009, the Wisconsin Alzheimer’s Disease Research Center (Wisconsin-ADRC) is one of the Alzheimer’s Disease Research Centers (ADRCs) designated by the National Institute on Aging (NIA) in the United States^2^. The Wisconsin ADRC conducts longitudinal follow-up of its participants. Eligible individuals were adults aged 45 years or older with decisional capacity and fluency in English. Exclusion criteria included having an active major medical or psychiatric illness or lacking a study partner.

Alzheimer’s Disease Neuroimaging Initiative: Launched in 2003 as a public–private partnership led by Principal Investigator Michael W. Weiner, MD, the Alzheimer’s Disease Neuroimaging Initiative (ADNI) seeks to determine whether serial MRI, PET imaging, other biomarkers, and clinical and neuropsychological assessments can be combined to monitor the progression of mild cognitive impairment (MCI) and early AD^3^. For the most current information, visit [www.adni-info.org](http://www.adni-info.org).

Washington Heights-Hamilton Heights-Inwood Columbia Aging Project (WHICAP): The Washington Heights–Inwood Columbia Aging Project (WHICAP) began in 1992 as a multi-ethnic, community-based prospective cohort study designed to investigate clinical and genetic risk factors for dementia^4^. Participants were enrolled in three waves (1992, 1999, and 2009) using consistent study protocols. Recruitment targeted older adults living in the northern Manhattan community. At baseline, participants completed a structured interview on general health and functional status, followed by a comprehensive assessment that included medical, neurological, and psychiatric histories, as well as standardized physical, neurological, and neuropsychological examinations. Follow-up visits occurred every 18–24 months using procedures consistent with those at baseline.

Estudio Familiar de Influencia Genética en Alzheimer: Initiated in 1998, the Estudio Familiar de Influencia Genética en Alzheimer (EFIGA) is a study of individuals of Caribbean Hispanic descent, including those with familial or sporadic late-onset AD as well as cognitively healthy, non-demented participants from the Dominican Republic and New York^5^. EFIGA collects detailed information on dementia status, general health, and functional abilities, using the same diagnostic criteria and protocols as WHICAP.

ROSMAP: The Religious Order Study and Memory and Aging Project (ROSMAP) integrates two longitudinal cohort studies that have been collecting detailed clinical, pathological, and molecular data since the early 1990s. The Religious Orders Study (ROS) and the Rush Memory and Aging Project (MAP) began in 1994 and 1997, respectively, and are collectively referred to as ROSMAP. Both studies are prospective cohort investigations of risk factors for cognitive decline, incident AD dementia, and other health outcomes. They share core design features of analytic cohort studies and require participants to consent to organ donation as a condition of enrollment. In addition to extensive clinicopathological data, ROSMAP includes a wide range of multi-omic data, providing a rich resource for studying molecular mechanisms underlying neurodegenerative disease and aging^6,7^.

HABS-HD: The Health and Aging Brain Study–Health Disparities (HABS-HD) is an ongoing community-based cohort study designed to examine factors related to brain health within the framework of AD biology. The study is grounded in the amyloid (A), tau (T), and neurodegeneration (N) [A/T(N)] framework, which was developed to characterize and stage the AD pathobiological cascade, and aims to evaluate its utility in more diverse populations^8^.

*Biomarker acquisition and processing*

Plasma biomarkers for WRAP and the Wisconsin ADRC were analyzed at the University of Gothenburg^9^. The primary biomarker, P-tau217, was measured in duplicate using the commercially available ALZpath assay (ALZpathDX, Carlsbad, CA), noted for its high sensitivity, precision, and reproducibility. All biomarker values were z-standardized relative to the AD-negative, cognitively unimpaired control group.

Plasma P-tau217 was measured using the validated, highly automated Lumipulse chemiluminescent enzyme immunoassay platform (G1200)^10,11^. Plasma data were downloaded from the ADNI database (https://ida.loni.usc.edu) in mid-2025.

In WHICAP and EFIGA, blood for plasma was collected in dipotassium ethylenediaminetetraacetic acid tubes and centrifuged at 2,000 g for 15 minutes at 4 °C. The resulting plasma was aliquoted into polypropylene tubes, frozen, and stored at –80 °C. Plasma P-tau217 was measured using the Simoa HD-X platform (Quanterix) with an assay specific to this biomarker. Prior to analysis, values more than five standard deviations above or below the sample mean were excluded^4,12,13^.

In ROSMAP, blood samples were collected during home visits into 2-mL EDTA tubes by trained phlebotomists or nurses experienced in venipuncture and transported to the Rush Alzheimer’s Disease Center laboratory. Following centrifugation, samples were aliquoted and stored at −80 °C. Plasma P-tau181 and P-tau217 concentrations were measured using immunoassays developed by Lilly Research Laboratories on the Meso Scale Discovery (MSD) platform, as previously described^14–16^.

In HABS-HD, plasma samples were assayed for P-tau217 using the Quanterix platform (AlzPath single-plex) in accordance with the manufacturer’s recommendations.

*Clinical diagnosis of AD/MCI*

In WRAP and Wisconsin-ADRC, diagnoses of AD and MCI are determined by a consensus review committee composed of dementia-specialist physicians, neuropsychologists, and nurse practitioners, following the National Institute on Aging–Alzheimer’s Association criteria^1,17^. MCI is diagnosed when the following criteria are met: (1) patient or informant concern regarding a change in cognition; (2) clear impairment in one or more cognitive domains; and (3) absence of criteria for dementia. In both WRAP and Wisconsin-ADRC, a subset of participants classified as ‘Impaired-other’ was excluded from the analysis.

In ADNI, diagnoses of AD and MCI are based on the Clinical Dementia Rating (CDR) score^18^. A global CDR score of 0 defined cognitively unimpaired status, while a score of 0.5 with a mandatory memory box score ≥0.5 defined MCI. Mild dementia was classified as a global CDR score of 0.5 or 1. In addition, ADNI criteria for MCI included: (1) subjective memory complaints reported by the participant, study partner, or clinician; (2) objective memory loss defined as scoring below an education-adjusted cut-off on delayed recall of Story A from the WMS-R Logical Memory Test (score ≤8 for ≥16 years of education; ≤4 for 8–15 years; ≤2 for 0–7 years); (3) global CDR score of 0.5; and (4) preserved general cognitive and functional performance such that a dementia diagnosis could not be made by the site physician at screening^19^.

In WHICAP and EFIGA, the final diagnosis of dementia in both cohorts was determined based on the criteria from the National Institute of Neurological and Communicative Disorders and Stroke–Alzheimer’s Disease and Related Disorders Association^20^. This determination occurred during diagnostic consensus conferences, which were attended by a panel of neurologists, psychiatrists, and neuropsychologists, using results from the neuropsychological battery and evidence of impairment in social or occupational functions. A diagnosis of MCI was made when cognitive impairment was present but did not meet the dementia criteria. Conversely, a cognitively intact diagnosis was assigned when no cognitive impairment was detected, or when the individual did not meet the criteria for dementia^21^.

In ROSMAP, participants received annual in-home clinical evaluations conducted by trained nurses and research staff. These evaluations included medical history interviews, medication review, blood collection, cognitive testing, and neurological examination. Alzheimer’s dementia diagnoses were made by a clinician using all available information, and were based on the 1984 McKhann criteria, which require evidence of cognitive decline with impairment in memory and at least one additional cognitive domain. Participants with cognitive impairment who did not meet criteria for dementia were classified as having MCI.^15^

In HABS-HD, cognitive diagnoses were determined through a consensus review process led by dementia experts affiliated with the study. An established diagnostic algorithm was consistent with standard diagnostic criteria for dementia. All participants classified as having MCI or dementia, as well as a random 10% sample of cognitively unimpaired participants, underwent consensus review that included independent chart review by dementia experts, a medical professional, and study clinic staff^8^.

*P-tau217 standardization and positivity classification*

For the baseline cross-sectional analysis, P‑tau217 values were log₁₀-transformed and z-standardized within each cohort to harmonize measurement scales across platforms, regardless of amyloid status. For longitudinal analyses, z-standardization was performed within each cohort using only individuals who were cognitively unimpaired at baseline. For analyses involving P‑tau217 positivity, we applied a three-range approach and defined two cohort-specific thresholds *a priori*: one that maximized sensitivity for identifying amyloid-positive AD and another that maximized specificity to reduce false positives (Supplementary Table 1)^2^. EFIGA, ROSMAP, and HABS-HD included multiple ancestries and did not generate cohort-specific thresholds. We used the WHICAP multi-ethnic threshold as a proxy. In the sensitivity analyses, we repeated the analyses in these three cohorts using the threshold defined by WRAP.

*Temporal dynamics of P‑tau217 in predicting cognitive impairment*

In the pooled sample and within each cohort, we conducted post hoc analyses to examine the temporal trajectory of plasma P‑tau217 in predicting cognitive impairment. We first estimated model-based restricted mean survival time (RMST) for remaining cognitively unimpaired over the entire follow-up period^22–25^. For analyses using the continuous, harmonized P-tau217 measure, we visualized RMST as a smooth function of P-tau217 to characterize dose-response patterns across its distribution. For binary analyses, RMST curves were estimated separately for P-tau217-positive and -negative groups. For the binary predictor, 95% confidence intervals were obtained via 1,000 bootstrap resamples. For the continuous predictor, confidence intervals were not displayed because the RMST curves were intended for descriptive visualization of the overall trajectory rather than formal pointwise inference. In the EFIGA cohort, the limited number of events precluded reliable estimation of confidence intervals. Second, using both the continuous and binary P-tau217 measures, we plotted covariate-adjusted survival curves stratified by P‑tau217 risk categories to identify the earliest time point at which survival probabilities began to diverge^26^. The proportional hazards assumption was violated in the combined sample for the P-tau217 positivity models due to heterogeneity across datasets; thus we applied a pseudo-value approach^24^. This method estimates survival at prespecified time points, fits generalized estimating equations to adjust for covariates, and averages predictions across groups to yield confounder-adjusted trajectories, with 1,000 bootstrap resamples for confidence intervals. When P-tau217 was modeled as continuous, a pseudo-value model was not available, therefore we stratified on the covariates that failed the proportional hazards test before plotting the survival curves. Pointwise bootstrap confidence intervals were calculated using the default number of 300 resamples. When the proportional hazards assumption was met, covariate-adjusted survival curves were derived directly from the fitted Cox model (e.g., cohort-specific analysis). In the pooled analyses, survival and RMST estimates were truncated at 14 years for comparability across strata because follow-up beyond this point was sparse in the smallest subsample (e.g., *APOE*-ε4 carriers), resulting in imprecise tail estimates; approximately 99% of observations occurred before 14 years.

All post hoc analyses were conducted in the full sample and stratified by *APOE*-ε4 status. All models were adjusted for baseline age, sex, education, *APOE* genotype (if applicable), cohort (if applicable), and ethnic group (if applicable). Analyses were conducted using complete cases; participants with missing data for *APOE* genotype, P‑tau217, clinical outcomes, or covariates were excluded.

*Differences in adjusted survival curves to identify the earliest point of divergence*

In the pooled sample and stratified by *APOE*-ε4 carrier status, we estimated risk-difference curves to determine the earliest time point at which two groups showed statistically significant difference in the covariate-adjusted survival curves^26,27^. Statistical testing was performed only at integer follow-up years. For continuous plasma P-tau217, risk differences were calculated by comparing adjusted survival probabilities at P-tau217 levels corresponding to +1 standard deviation above the mean versus the mean value. For binary P-tau217 positivity, we compared adjusted survival probabilities between P-tau217-positive and P-tau217-negative individuals. The risk difference was defined as the difference between the two adjusted survival probability estimates for the specified risk groups. The standard error (SE) of the difference was computed using the SE of the two probability estimates. Confidence intervals were calculated using this SE and the normal approximation. P-values were derived using the same SE in a two-sided one-sample t-test, with the null hypothesis that the difference equals zero. To account for multiple testing across time points, we applied the Benjamini–Yekutieli false discovery rate (BY-FDR) procedure, which is robust to dependence among tests.

*Time to 75% cognitive-impairment-free survival*

Based on the covariate-adjusted survival curves, we estimated the confounder-adjusted time to 75% cognitive-impairment-free survival (S(t)=0.75) and calculated the difference between the corresponding survival time quantiles across groups^27–29^. Time to 75% cognitive-impairment–free survival was defined as the time at which 75% of participants were expected to remain cognitively unimpaired (i.e., 25% cumulative incidence of impairment). We selected the 75% cognitive-impairment-free survival threshold because several cohorts were relatively young and experienced few events (e.g., Wisconsin), so the median survival time could not be estimated for some risk groups. In contrast, the time at which the adjusted survival probability reached 0.75 could be consistently estimated across cohorts. The survival time-quantile difference was defined as the horizontal separation between the two adjusted survival curves at the same survival probability level and was obtained by comparing the times at which each curve reached S(t)=0.75. For categorical analyses, confidence intervals were estimated because the survival time-quantile difference was defined for a prespecified comparison between two discrete groups. For the continuous predictor, the survival time quantile was plotted as a smooth function of P-tau217 across its distribution to illustrate the overall descriptive trajectory rather than support pointwise inference at individual biomarker values; confidence intervals were therefore not presented. For categorical analyses, confidence intervals were estimated using a bootstrap procedure. For each bootstrap resample, the adjusted survival curve and corresponding survival time quantile were recalculated, and the SE for each group was obtained as the standard deviation of the bootstrap-derived quantile estimates. For between-group comparisons, the SE of the difference was computed using the pooled SE of the two group-specific estimates and used with a normal approximation to construct confidence intervals. P-values were calculated using a two-sided one-sample t-test with the null hypothesis that the difference in survival time quantiles equals zero.

*Accelerated failure time (AFT) models*

We used an accelerated failure time (AFT) model to evaluate the joint effect of plasma P-tau217 and *APOE*-ε4 status on time to cognitive impairment in the pooled sample. The AFT model was chosen because it directly models survival time and does not rely on the proportional hazards assumption required by the Cox proportional hazards model, which instead models covariate effects on the hazard function^30^. In the AFT model, regression coefficients are interpreted using the time ratio (TR), calculated as exp(β). A TR < 1 indicates a shorter event-free interval (earlier occurrence of cognitive impairment), TR = 1 indicates no effect on survival time, and TR > 1 indicates a longer event-free interval (delayed cognitive impairment). Thus, covariates in the AFT model are interpreted as accelerating or decelerating the timing of clinical onset. We also performed AFT model analyses within each cohort; cohort-specific results are available upon request.

*Association of APOE with cases defined by both clinical cognitive impairment and P-tau217 positivity*

To assess the effects of *APOE* across the full AD continuum, we examined *APOE* associations with case groups defined jointly by clinical cognitive status and plasma P-tau217 positivity. Participants were classified into four mutually exclusive groups: (1) P-tau217-positive with clinical cognitive impairment, representing AD-related cognitive impairment; (2) P-tau217-positive without clinical impairment, representing the preclinical phase of AD; (3) P-tau217-negative with clinical impairment, suggesting cognitive impairment due to non-AD pathology; and (4) P-tau217-negative without clinical impairment, representing cognitively normal controls. For all *APOE* association analyses, the P-tau217-negative cognitively normal group was used as the reference category. Because established P-tau217 cutoffs were not available in EFIGA, ROSMAP, and HABS-HD, we applied the WHICAP cutoff as a proxy, as these cohorts are similarly multi-ancestry and predominantly non-White populations. These analyses were conducted cross-sectionally at baseline only, because longitudinal P-tau217 measurements were not available for most included cohorts. Analyses were performed in the pooled sample and separately within each cohort; cohort-specific results are available upon request.

*Random forest prediction*

We used a non-parametric random survival forest (RSF) model to predict future cognitive impairment based on baseline plasma P-tau217, *APOE*-ε4 carrier status, and additional covariates^31^. Continuous P-tau217 was used because continuous predictors are better suited for random forest models than dichotomized variables. The covariates included age, sex, education, cohort, and ethnicity. Model performance was evaluated using a leave-one-cohort-out validation framework. In each iteration, all cohorts except one were pooled to train the model, and the remaining cohort was used for external validation. This procedure was repeated for each cohort except HABS-HD and EFIGA, which had limited follow-up time (these cohorts were included in the training data but not used as validation cohorts). For each validation cohort, predictive performance was assessed using the time-dependent area under the curve (time-dependent AUC). We evaluated several models which include *APOE*-ε4 with covariates, P-tau217 with covariates, P-tau217 plus *APOE*-ε4 with covariates, and P-tau217 models fitted separately among ε4 carriers and non-carriers with covariates. The RSF model was implemented using 1,000 trees with a minimum terminal node size of 5 individuals. At each node, candidate split points were evaluated from 50 randomly sampled cut points. All baseline predictors were included without prior variable selection.

*Other methods*

In cohort-specific longitudinal analyses, models from cohorts with small sample size (e.g., EFIGA) did not adjust for *APOE* genotype because estimates were unstable due to few ε4/ε4 carriers, which affected subsequent survival curve plotting.

*R Packages included in the analyses*

Survival analyses: survival (3.7.0), survminer (0.5.0), (adjustedCurves) 0.11.2, (contsurvplot) 0.2.2

Meta analysis: meta (8.0.2), metafor (4.8.0), timeROC (0.4)

Random forest: randomForestSRC (3.0)

Plotting: ggplot2 (3.5.1)

**Supplementary Table 1. Cutoffs used in the analyses for each cohort.**

| **Cohort** | **Proxy cutoff?** | **Cut-off maximizing sensitivity** | **Cut-off maximizing specificity** |
| --- | --- | --- | --- |
| Wisconsin | No | P-tau217 > 0.40 | P-tau217 > 0.63 |
| ADNI | No | P-tau217 > 0.167 | P-tau217 > 0.334 |
| EFIGA | Yes | P-tau217 > 0.40675 | P-tau217 > 0.81 |
| WHICAP | No | P-tau217 > 0.40675 | P-tau217 > 0.81 |
| ROSMAP | Yes | P-tau217 > 0.40675 | P-tau217 > 0.81 |
| HABS-HD | Yes | P-tau217 > 0.40675 | P-tau217 > 0.81 |

**Supplementary Table 1** presents the cutoffs used to define plasma P-tau217 positivity in the current analyses. Cutoffs for each cohort were obtained from published studies or technical handbooks. Because established cutoffs were not available for EFIGA, ROSMAP, and HABS-HD, we applied the WHICAP cutoff as a proxy, given that these cohorts are similarly multi-ancestry and predominantly non-White populations.

**Supplementary Table 2. Baseline characteristics of a subset of cognitive unimpaired participants included in the longitudinal analysis**

| **Characteristic** | **ADNI** N = 401*^1^* | **EFIGA** N = 329*^1^* | **HABS-HD** N = 1,169*^1^* | **ROSMAP** N = 732*^1^* | **WHICAP** N = 800*^1^* | **Wisconsin** N = 1,138*^1^* |
| --- | --- | --- | --- | --- | --- | --- |
| **Age at baseline (years)** | 73 (7) | 69 (7) | 66 (8) | 78 (7) | 75 (6) | 62 (7) |
| **Sex** |  |  |  |  |  |  |
| Women | 228 (57%) | 237 (72%) | 777 (66%) | 600 (82%) | 554 (69%) | 777 (68%) |
| Men | 173 (43%) | 92 (28%) | 392 (34%) | 132 (18%) | 246 (31%) | 361 (32%) |
| **Education (years)** | 17 (2) | 7 (5) | 14 (4) | 16 (4) | 12 (5) | 16 (2) |
| **Follow-up years** | 4.03 (2.39) | 2.77 (1.05) | 3.42 (1.64) | 5.65 (4.01) | 6.23 (3.21) | 6.40 (2.94) |
| **Initial P-tau217 (pg/mL)** | 0.19 (0.19) | 0.35 (0.23) | 0.39 (0.29) | 0.60 (0.60) | 0.43 (0.33) | 0.35 (0.22) |
| **Initial P-tau217 positivity (sensitivity-cutoff)** | | | | | | |
| Negative | 262 (65%) | 246 (75%) | 829 (71%) | 317 (43%) | 531 (66%) | 843 (74%) |
| Positive | 139 (35%) | 83 (25%) | 340 (29%) | 415 (57%) | 269 (34%) | 295 (26%) |
| **Initial P-tau217 positivity (specificity-cutoff)** | | | | | | |
| Negative | 351 (88%) | 321 (98%) | 1,085 (93%) | 597 (82%) | 734 (92%) | 1,039 (91%) |
| Positive | 50 (12%) | 8 (2.4%) | 84 (7.2%) | 135 (18%) | 66 (8.3%) | 99 (8.7%) |
| **Diagnosis at censoring or event^2^** | | | | | | |
| Incident impairment | 146 (36%) | 43 (13%) | 172 (15%) | 340 (46%) | 342 (43%) | 65 (5.7%) |
| Unimpaired | 255 (64%) | 286 (87%) | 997 (85%) | 392 (54%) | 458 (57%) | 1,073 (94%) |
| **Ethnic group^3^** |  |  |  |  |  |  |
| Black | 52 (13%) | 0 (0%) | 160 (14%) | 285 (39%) | 212 (27%) | 36 (3.2%) |
| Non-Hispanic White | 319 (80%) | 0 (0%) | 564 (48%) | 385 (53%) | 239 (30%) | 1,065 (94%) |
| Others and Hispanics | 30 (7.5%) | 329 (100%) | 445 (38%) | 62 (8.5%) | 349 (44%) | 37 (3.3%) |
| ***APOE*^4^** |  |  |  |  |  |  |
| 2/3 | 39 (9.7%) | 40 (12%) | 120 (10%) | 107 (15%) | 114 (14%) | 101 (8.9%) |
| 3/3 | 216 (54%) | 192 (58%) | 770 (66%) | 446 (61%) | 491 (61%) | 626 (55%) |
| 3/4 | 131 (33%) | 92 (28%) | 258 (22%) | 165 (23%) | 180 (23%) | 368 (32%) |
| 4/4 | 15 (3.7%) | 5 (1.5%) | 21 (1.8%) | 14 (1.9%) | 15 (1.9%) | 43 (3.8%) |
| *^1^* Mean (SD); n (%) | | | | | | |

**Supplementary Table 2** presents summary statistics for baseline characteristics of a subset of cognitively unimpaired participants included in the longitudinal analyses across cohorts. Biomarker availability and longitudinal follow-up varied by cohort; the baseline sample reflects the first available plasma biomarker measurement for each participant.

^2^ Baseline cognitive status was defined at the time when plasma biomarker data first became available for each participant.

^3^ Ethnic group identity was collected in detail within each cohort; however, the distribution varied substantially across studies. For example, Wisconsin and ADNI were predominantly composed of non-Hispanic White participants, followed by Black individuals, with very few Hispanic or other ethnic groups. In contrast, EFIGA included only Hispanic participants, while WHICAP had a more balanced ethnic composition. To facilitate analysis and ensure adequate sample sizes within each category, we combined Hispanic and other ethnic groups into a single category. This grouping also helped stabilize regression models by reducing sparse categories and improving model convergence.

^4^ *APOE* ε2/ε2 and ε2/ε4 genotypes were excluded due to their low frequency and the potentially protective effects of the ε2 allele, which may confound interpretation of *APOE*-ε4–related risk associations.

**Supplementary Table 3**. Interaction p-values for P-tau217 × *APOE*-ε4 in Cox proportional hazards models for the combined sample and individual cohorts

|  | **Continues P-tau217**  **(log-z)** | **P-tau217 positivity**  **(sensitivity cutoff)** | **P-tau217 positivity**  **(specificity cutoff)** |
| --- | --- | --- | --- |
| Wisconsin | **0.001** | **0.004** | 0.069 |
| ADNI | **0.005** | 0.213 | 0.163 |
| EFIGA | 0.322 | 0.870 | 0.933 |
| WHICAP | 0.151 | 0.292 | 0.952 |
| ROSMAP | 0.637 | 0.621 | 0.443 |
| HABS-HD | 0.324 | 0.886 | 0.541 |
| Combine | **<0.001** | **0.005** | **0.011** |

**Supplementary Table 3** presents the interaction p-values for P-tau217 × *APOE*-ε4 from Cox proportional hazards models in the combined sample and individual cohorts, with P-tau217 modeled as a continuous variable and as a binary variable using cutoffs that maximized sensitivity and specificity. Because established cutoffs were not available for EFIGA, ROSMAP, and HABS-HD, we applied the WHICAP cutoff as a proxy, given that these cohorts are similarly multi-ancestry and predominantly non-White populations. Continuous P-tau217 values were harmonized using log₁₀ transformation followed by z-standardization. All interaction tests were derived from Cox proportional hazards models with incident cognitive impairment (i.e., MCI or AD) as the outcome, adjusted for age, sex, education, cohort (if applicable), and ethnicity. Bolded values indicate interaction p-values < 0.05.

**Supplementary Table 4. Accelerated failure time (AFT) models of time to cognitive impairment showing the *APOE-*ε4 × P-tau217 interaction in the pooled cohort**

| **Model terms** | **Beta** | **SE** | **Time Ratio** | **Lower CI** | **Upper CI** | **P-value** |
| --- | --- | --- | --- | --- | --- | --- |
| *Continuous P-tau217* |  |  |  |  |  |  |
| *APOE*-ε4 | -0.04 | 0.05 | 0.96 | 0.88 | 1.05 | 3.29E-01 |
| P-tau217 (log-z) | -0.13 | 0.02 | 0.87 | 0.84 | 0.91 | **5.16E-09** |
| *APOE*-ε4 × P-tau217 (log-z) | -0.12 | 0.04 | 0.89 | 0.82 | 0.95 | **1.13E-03** |
| *Sensitivity cutoff* |  |  |  |  |  |  |
| *APOE*-ε4 | 0.01 | 0.06 | 1.01 | 0.89 | 1.14 | 9.27E-01 |
| P-tau217 positivity | -0.18 | 0.05 | 0.84 | 0.77 | 0.91 | **8.63E-05** |
| *APOE*-ε4 × P-tau217 positivity | -0.24 | 0.08 | 0.79 | 0.67 | 0.93 | **3.87E-03** |
| *Specificity cutoff* |  |  |  |  |  |  |
| *APOE*-ε4 | -0.07 | 0.05 | 0.94 | 0.85 | 1.03 | 1.58E-01 |
| P-tau217 positivity | -0.41 | 0.06 | 0.66 | 0.59 | 0.75 | **3.09E-11** |
| *APOE*-ε4 × P-tau217 positivity | -0.21 | 0.09 | 0.81 | 0.68 | 0.97 | **2.16E-02** |

**Supplementary Table 4** shows AFT models of time to cognitive impairment including the *APOE*-ε4 × P-tau217 interaction in the pooled cohort. The time ratio (TR) reflects acceleration or delay in time to the event by a multiplicative factor (e.g., TR = 0.87 indicates a 13% shorter time to cognitive impairment). P-tau217 positivity was defined using cohort-specific thresholds that maximized sensitivity or specificity. Because established cutoffs were not available for EFIGA, ROSMAP, and HABS-HD, we applied the WHICAP cutoff as a proxy, given that these cohorts are similarly multi-ancestry and predominantly non-White populations. Additional covariates include age, sex, education, ethnicity, and cohort. Bolded values indicate significance at p < 0.05.

**Supplementary Table 5.** Association of *APOE* with case groups defined by clinical cognitive impairment and P-tau217 positivity based on cutoffs maximizing sensitivity and specificity in the pooled sample and by ancestry at baseline

A. *P-tau217 positivity determined by the cutoff maximizing sensitivity*

|  | **All sample** | | | | **Non-Hispanic White** | | | | **Black** | | | | **Hispanic and others** | | | |
| --- | --- | --- | --- | --- | --- | --- | --- | --- | --- | --- | --- | --- | --- | --- | --- | --- |
| ***APOE*** | **Case** | **Control** | **OR (CI)** | **P-value** | **Case** | **Control** | **OR (CI)** | **P-value** | **Case** | **Control** | **OR (CI)** | **P-value** | **Case** | **Control** | **OR (CI)** | **P-value** |
| **Clinical cognitive impairment & P-tau217 positive (sensitivity-cutoff) vs clinical cognitive unimpaired & P-tau217 negative** | | | | | | | | | | | | | | | | |
| ε4 vs non-ε4 | 1286 | 4299 | **4.57 (3.85, 5.42)** | **2.97E-68** | 602 | 2060 | **7.12 (5.36, 9.45)** | **5.56E-42** | 205 | 671 | **2.43 (1.66, 3.55)** | **4.56E-06** | 479 | 1568 | **3.98 (3.03, 5.21)** | **1.39E-23** |
| ε2/ε3 vs ε3/ε3 | 1286 | 4299 | **0.65 (0.49, 0.87)** | **4.13E-03** | 602 | 2060 | **0.42 (0.25, 0.70)** | **8.08E-04** | 205 | 671 | 0.80 (0.44, 1.46) | 4.61E-01 | 479 | 1568 | 0.77 (0.48, 1.22) | 2.66E-01 |
| ε3/ε4 vs ε3/ε3 | 1286 | 4299 | **3.46 (2.89, 4.16)** | **2.22E-40** | 602 | 2060 | **4.93 (3.66, 6.65)** | **1.60E-25** | 205 | 671 | **1.91 (1.25, 2.90)** | **2.68E-03** | 479 | 1568 | **3.24 (2.42, 4.33)** | **2.09E-15** |
| ε4/ε4 vs ε3/ε3 | 1286 | 4299 | **17.19 (11.84, 24.95)** | **1.31E-50** | 602 | 2060 | **39.74 (21.39, 73.84)** | **2.26E-31** | 205 | 671 | **6.60 (3.07, 14.16)** | **1.32E-06** | 479 | 1568 | **11.82 (6.43, 21.71)** | **1.75E-15** |
| **Clinical cognitive unimpaired & P-tau217 positive (sensitivity-cutoff) vs clinical cognitive unimpaired & P-tau217 negative** | | | | | | | | | | | | | | | | |
| ε4 vs non-ε4 | 2016 | 4299 | **2.59 (2.28, 2.93)** | **5.13E-49** | 1181 | 2060 | **3.08 (2.59, 3.66)** | **2.63E-37** | 317 | 671 | **1.50 (1.10, 2.05)** | **1.12E-02** | 518 | 1568 | **2.54 (2.01, 3.22)** | **1.05E-14** |
| ε2/ε3 vs ε3/ε3 | 2016 | 4299 | **0.78 (0.65, 0.95)** | **1.55E-02** | 1181 | 2060 | **0.69 (0.52, 0.91)** | **7.77E-03** | 317 | 671 | 0.96 (0.61, 1.51) | 8.61E-01 | 518 | 1568 | 0.84 (0.58, 1.22) | 3.61E-01 |
| ε3/ε4 vs ε3/ε3 | 2016 | 4299 | **2.33 (2.04, 2.67)** | **2.81E-35** | 1181 | 2060 | **2.70 (2.25, 3.24)** | **6.90E-27** | 317 | 671 | 1.35 (0.96, 1.90) | 8.26E-02 | 518 | 1568 | **2.41 (1.87, 3.09)** | **5.60E-12** |
| ε4/ε4 vs ε3/ε3 | 2016 | 4299 | **4.69 (3.37, 6.52)** | **4.05E-20** | 1181 | 2060 | **6.55 (4.11, 10.44)** | **3.01E-15** | 317 | 671 | **3.12 (1.47, 6.60)** | **3.01E-03** | 518 | 1568 | **3.35 (1.78, 6.34)** | **1.91E-04** |
| **Clinical cognitive impairment & P-tau217 negative (sensitivity-cutoff) vs clinical cognitive unimpaired & P-tau217 negative** | | | | | | | | | | | | | | | | |
| ε4 vs non-ε4 | 981 | 4299 | 1.09 (0.91, 1.31) | 3.47E-01 | 213 | 2060 | 0.84 (0.56, 1.24) | 3.74E-01 | 187 | 671 | 0.87 (0.59, 1.26) | 4.50E-01 | 581 | 1568 | **1.30 (1.01, 1.68)** | **3.84E-02** |
| ε2/ε3 vs ε3/ε3 | 981 | 4299 | 0.98 (0.78, 1.23) | 8.56E-01 | 213 | 2060 | 0.83 (0.52, 1.33) | 4.36E-01 | 187 | 671 | 0.95 (0.58, 1.56) | 8.33E-01 | 581 | 1568 | 1.08 (0.78, 1.49) | 6.53E-01 |
| ε3/ε4 vs ε3/ε3 | 981 | 4299 | 1.06 (0.87, 1.28) | 5.62E-01 | 213 | 2060 | 0.77 (0.51, 1.16) | 2.15E-01 | 187 | 671 | 0.81 (0.54, 1.22) | 3.11E-01 | 581 | 1568 | **1.31 (1.01, 1.71)** | **4.35E-02** |
| ε4/ε4 vs ε3/ε3 | 981 | 4299 | 1.46 (0.86, 2.50) | 1.65E-01 | 213 | 2060 | 1.68 (0.50, 5.59) | 4.00E-01 | 187 | 671 | 1.25 (0.52, 3.02) | 6.16E-01 | 581 | 1568 | 1.42 (0.63, 3.21) | 4.03E-01 |

B. *P-tau217 positivity determined by the cutoff maximizing specificity*

|  | **All sample** | | | | **Non-Hispanic White** | | | | **Black** | | | | **Hispanic and others** | | | |
| --- | --- | --- | --- | --- | --- | --- | --- | --- | --- | --- | --- | --- | --- | --- | --- | --- |
| ***APOE*** | **Case** | **Control** | **OR (CI)** | **P-value** | **Case** | **Control** | **OR (CI)** | **P-value** | **Case** | **Control** | **OR (CI)** | **P-value** | **Case** | **Control** | **OR (CI)** | **P-value** |
| **Clinical cognitive impairment & P-tau217 positive (specificity-cutoff) vs clinical cognitive unimpaired & P-tau217 negative** | | | | | | | | | | | | | | | | |
| ε4 vs non-ε4 | 735 | 5740 | **5.02 (4.16, 6.07)** | **8.80E-63** | 413 | 2844 | **6.04 (4.60, 7.94)** | **5.60E-38** | 93 | 906 | **3.76 (2.30, 6.16)** | **1.43E-07** | 229 | 1990 | **4.25 (3.10, 5.84)** | **3.83E-19** |
| ε2/ε3 vs ε3/ε3 | 735 | 5740 | **0.53 (0.36, 0.79)** | **1.73E-03** | 413 | 2844 | **0.45 (0.25, 0.79)** | **5.92E-03** | 93 | 906 | 0.62 (0.25, 1.53) | 2.96E-01 | 229 | 1990 | 0.59 (0.29, 1.19) | 1.42E-01 |
| ε3/ε4 vs ε3/ε3 | 735 | 5740 | **3.73 (3.04, 4.57)** | **1.25E-36** | 413 | 2844 | **4.37 (3.26, 5.85)** | **3.70E-23** | 93 | 906 | **2.55 (1.46, 4.43)** | **9.52E-04** | 229 | 1990 | **3.34 (2.37, 4.71)** | **4.75E-12** |
| ε4/ε4 vs ε3/ε3 | 735 | 5740 | **18.25 (12.65, 26.32)** | **1.98E-54** | 413 | 2844 | **25.27 (14.70, 43.46)** | **1.64E-31** | 93 | 906 | **13.04 (5.65, 30.11)** | **1.80E-09** | 229 | 1990 | **13.05 (6.90, 24.67)** | **2.77E-15** |
| **Clinical cognitive unimpaired & P-tau217 positive (specificity-cutoff) vs clinical cognitive unimpaired & P-tau217 negative** | | | | | | | | | | | | | | | | |
| ε4 vs non-ε4 | 575 | 5740 | **3.43 (2.84, 4.14)** | **1.43E-37** | 397 | 2844 | **3.79 (3.00, 4.78)** | **3.10E-29** | 82 | 906 | **2.20 (1.36, 3.57)** | **1.34E-03** | 96 | 1990 | **3.41 (2.20, 5.31)** | **4.84E-08** |
| ε2/ε3 vs ε3/ε3 | 575 | 5740 | **0.57 (0.39, 0.83)** | **3.45E-03** | 397 | 2844 | **0.54 (0.34, 0.86)** | **9.60E-03** | 82 | 906 | 0.49 (0.20, 1.24) | 1.31E-01 | 96 | 1990 | 0.86 (0.37, 1.98) | 7.26E-01 |
| ε3/ε4 vs ε3/ε3 | 575 | 5740 | **2.88 (2.35, 3.52)** | **5.49E-25** | 397 | 2844 | **3.21 (2.51, 4.11)** | **1.32E-20** | 82 | 906 | 1.64 (0.97, 2.79) | 6.50E-02 | 96 | 1990 | **2.98 (1.85, 4.80)** | **6.63E-06** |
| ε4/ε4 vs ε3/ε3 | 575 | 5740 | **7.16 (4.80, 10.68)** | **5.18E-22** | 397 | 2844 | **7.05 (4.25, 11.68)** | **3.69E-14** | 82 | 906 | **5.96 (2.33, 15.21)** | **1.89E-04** | 96 | 1990 | **8.76 (3.48, 22.03)** | **4.01E-06** |
| **Clinical cognitive impairment & P-tau217 negative (specificity-cutoff) vs clinical cognitive unimpaired & P-tau217 negative** | | | | | | | | | | | | | | | | |
| ε4 vs non-ε4 | 1532 | 5740 | **1.23 (1.07, 1.42)** | **3.54E-03** | 402 | 2844 | 1.13 (0.87, 1.48) | 3.51E-01 | 299 | 906 | 1.03 (0.77, 1.39) | 8.23E-01 | 831 | 1990 | **1.38 (1.13, 1.70)** | **1.87E-03** |
| ε2/ε3 vs ε3/ε3 | 1532 | 5740 | 0.91 (0.75, 1.10) | 3.29E-01 | 402 | 2844 | 0.72 (0.49, 1.05) | 8.88E-02 | 299 | 906 | 0.91 (0.61, 1.37) | 6.50E-01 | 831 | 1990 | 1.02 (0.77, 1.36) | 8.89E-01 |
| ε3/ε4 vs ε3/ε3 | 1532 | 5740 | 1.13 (0.97, 1.32) | 1.05E-01 | 402 | 2844 | 0.94 (0.70, 1.25) | 6.48E-01 | 299 | 906 | 0.96 (0.69, 1.32) | 7.83E-01 | 831 | 1990 | **1.33 (1.07, 1.66)** | **9.11E-03** |
| ε4/ε4 vs ε3/ε3 | 1532 | 5740 | **2.17 (1.52, 3.10)** | **2.28E-05** | 402 | 2844 | **3.21 (1.73, 5.96)** | **2.15E-04** | 299 | 906 | 1.52 (0.76, 3.06) | 2.38E-01 | 831 | 1990 | **2.07 (1.16, 3.68)** | **1.32E-02** |

**Supplementary Table 5** presents the association of *APOE* with case groups defined jointly by clinical cognitive impairment and P-tau217 positivity based on cutoffs maximizing sensitivity and specificity in the pooled sample and by ancestry at baseline. We defined four groups according to P-tau217 status and clinical symptoms: (1) P-tau217-positive with clinical cognitive impairment, representing AD-related cognitive impairment; (2) P-tau217-positive without clinical impairment, representing the preclinical phase; (3) P-tau217-negative with clinical impairment, suggesting cognitive impairment due to non-AD pathology; and (4) P-tau217-negative without clinical impairment, representing healthy controls. For all *APOE* association analyses, the healthy control group was used as the reference group. Because established cutoffs were not available for EFIGA, ROSMAP, and HABS-HD, we applied the WHICAP cutoff as a proxy, given that these cohorts are similarly multi-ancestry and predominantly non-White populations. Additional covariates include age, sex, education, ethnicity (if applicable), and cohort. Bolded values indicate significance at p < 0.05.

**Supplementary Figure 1. Cohort-specific and meta-analytic hazard ratios for incident cognitive impairment in the full sample and *APOE*-stratified subgroups using binary P‑tau217 measures, along with adjusted survival probabilities illustrating the temporal dynamics of P-tau217 in the pooled cohort.**

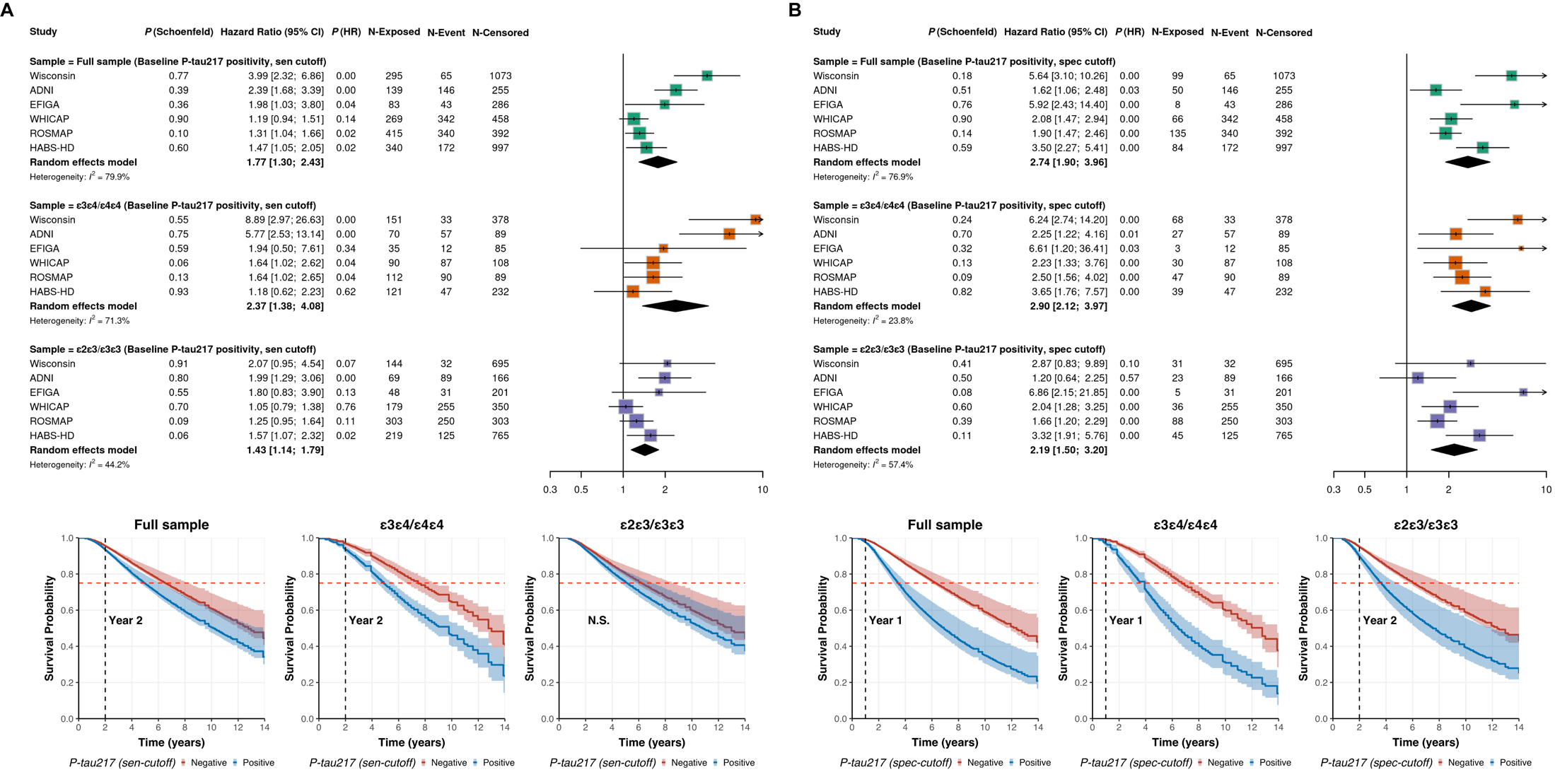

**Supplementary Figure 1** presents cohort-specific hazard ratios for incident cognitive impairment in the full sample and *APOE*-stratified subgroups, using binary P‑tau217 measures, along with meta-analytic estimates and adjusted survival probabilities illustrating the temporal dynamics of P-tau217 in the pooled cohort. Supplementary Figure 1A shows results when P-tau217 positivity is defined using the sensitivity-based cut-off, and Supplementary Figure 1B presents results when positivity is defined using the specificity-based cut-off. Given variability in longitudinal biomarker availability across cohorts, a standardized longitudinal analysis framework was applied. For each participant, baseline was defined as the time of first available biomarker measurement. In cohorts with only cross-sectional biomarker data, this represents a single time point; in longitudinal cohorts, it corresponds to the earliest biomarker collection. Clinical follow-up data were linked to each participant’s biomarker baseline, and only visits occurring after this time point were included. Analyses were restricted to individuals who were cognitively unimpaired at baseline and had follow-up data.

Incident cognitive impairment was defined as conversion from cognitively unimpaired to either MCI or AD. Time-to-event was defined as the interval from baseline to the date of diagnosis for converters or to the last available follow-up for censored cases. Cox proportional hazards models were used, adjusting for age, sex, education, ethnicity, and *APOE* genotype. The proportional hazards assumption was tested using Schoenfeld residuals. Within each full or stratified subgroup, random-effects meta-analyses were conducted using cohort-specific estimates. P‑tau217 positivity was defined using cohort-specific thresholds that maximized sensitivity or specificity. Because established cutoffs were not available for EFIGA, ROSMAP, and HABS-HD, we applied the WHICAP cutoff as a proxy, given that these cohorts are similarly multi-ancestry and predominantly non-White populations. P(Schoenfeld) indicates the p-value from the Schoenfeld global test for proportional hazards.

For the adjusted survival curve and to maximize statistical power, data from all cohorts were combined. P-tau217 positivity was defined within each cohort using thresholds that maximized sensitivity or specificity, then pooled across cohorts. Because the proportional hazards assumption was violated in the combined sample, we applied a pseudo-value approach (adjustedCurves package in R) when modelling P-tau217 as a binary positivity measure to address this issue. This method estimates survival probabilities at prespecified time points, adjusts for covariates (age, sex, education, cohort, ethnicity, and *APOE* genotype), and averages predictions across groups. In cohorts with small sample size (e.g., EFIGA), association models did not adjust for *APOE* genotype. Confidence intervals were obtained from 1,000 bootstrap resamples.

**Supplementary Figure 2. Model-based restricted mean survival time (RMST) curves showing time spent cognitively unimpaired by baseline P-tau217 positivity, defined using a cutoff that maximizes sensitivity and specificity, as well as by continuous P-tau217 levels, in the combined sample.**

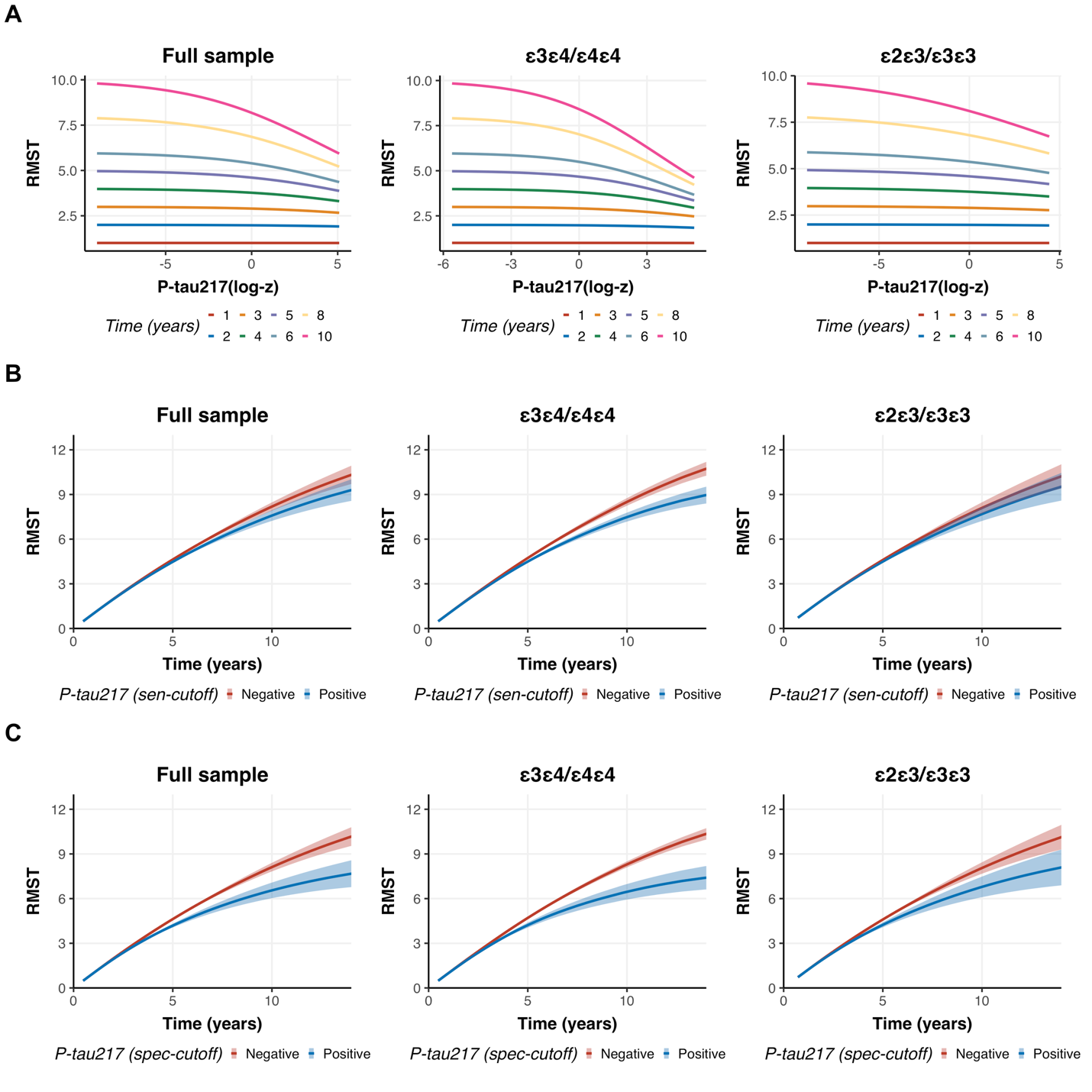

**Supplementary Figure 2** shows model-based restricted mean survival time (RMST) curves for time spent cognitively unimpaired, presented for the full sample and stratified by *APOE*-ε4 status, using baseline continuous P-tau217 levels (A) and binary P-tau217 positivity thresholds that maximize sensitivity (B) and specificity (C) as predictors. Because established cutoffs were not available for EFIGA, ROSMAP, and HABS-HD, we applied the WHICAP cutoff as a proxy, given that these cohorts are similarly multi-ancestry and predominantly non-White populations. Continuous P-tau217 values were harmonized using log₁₀ transformation followed by z-standardization within each cohort. RMST curves represent the estimated average duration participants remained cognitively unimpaired and were derived from Cox proportional hazards models with incident cognitive impairment (i.e., MCI or AD) as the outcome. All models were adjusted for age, sex, education, ethnicity, cohort, and *APOE* genotype. For continuous P-tau217, the x-axis represents harmonized continuous P-tau217 levels, and each curve corresponds to a specific follow-up year. The y-axis indicates RMST across baseline P-tau217 values. Flat (horizontal) curves at early follow-up years reflect minimal differences in RMST between individuals with low versus high P-tau217. At longer follow-up durations, curves slope downward, indicating shorter cognitively unimpaired periods for those with higher baseline P-tau217. For binary P-tau217 measures, the x-axis represents follow-up time (years), and the y-axis shows RMST, defined as the average time participants remained cognitively unimpaired up to each follow-up time point for each P-tau217 group. Overlapping curves indicate similar RMST estimates between groups at a given time point. Confidence intervals were estimated using 1,000 bootstrap resamples.

**Supplementary Figure 3. Adjusted survival probabilities illustrating the temporal dynamics of continuous plasma P-tau217 in predicting cognitive impairment across cohorts, in the full sample and stratified by *APOE*-ε4 status.**

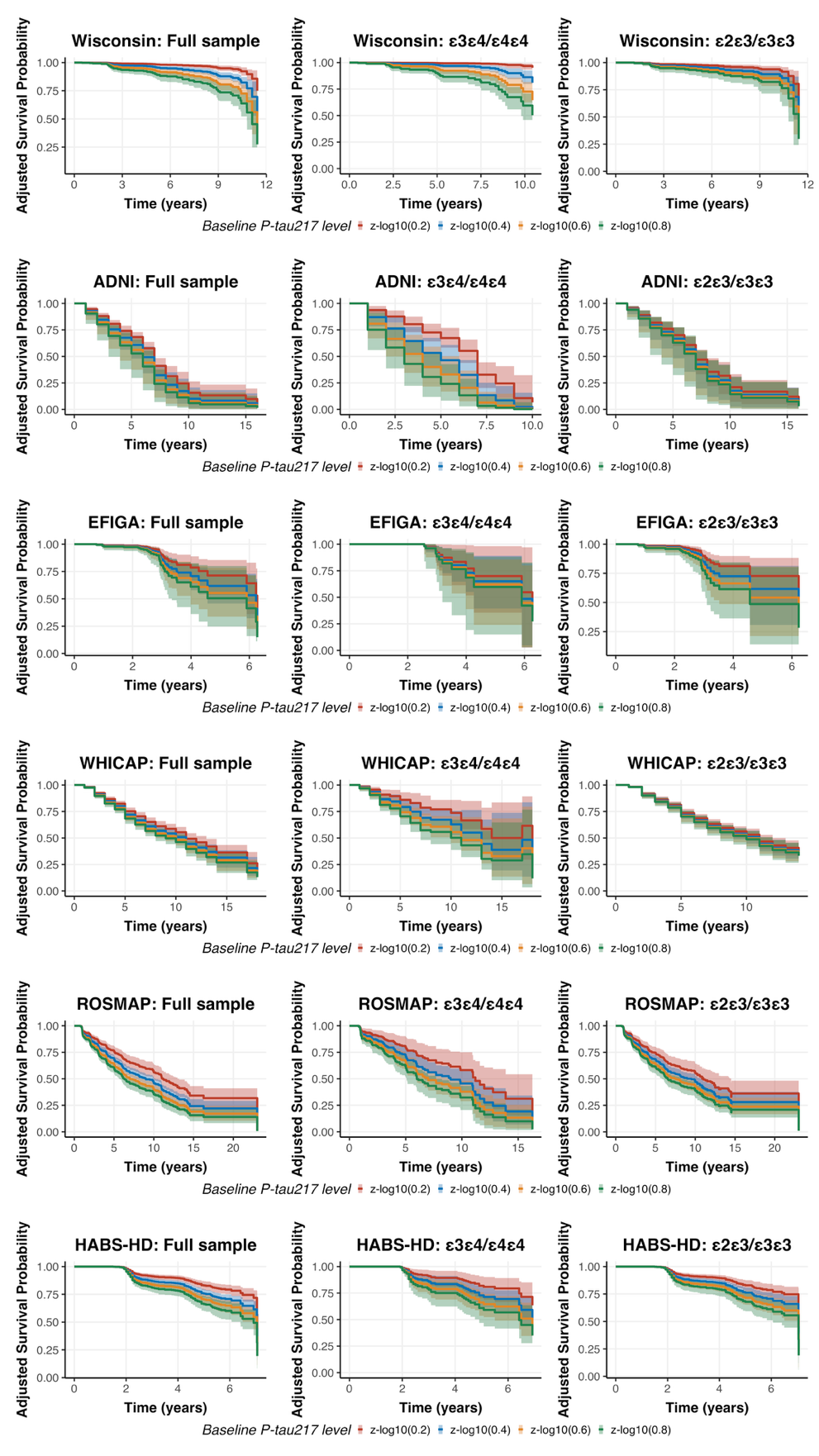

**Supplementary Figure 3** presents cohort-specific model-based curves of adjusted survival probability for cognitive impairment, shown for the full sample and stratified by *APOE*-ε4 status, using baseline continuous P-tau217 as the predictor. Continuous P-tau217 values were harmonized across cohorts using log₁₀ transformation followed by z-standardization. The adjusted survival probability curves represent the estimated probability of remaining cognitively unimpaired over time, after covariate adjustment, and were derived from Cox proportional hazards models with incident cognitive impairment (i.e., MCI or AD) as the outcome. All models were adjusted for age, sex, education, ethnicity, and *APOE* genotype. Stratification was applied in models where the proportional hazards assumption was violated. Within each cohort, the x-axis represents follow-up time (years), and the y-axis shows adjusted survival probability for individuals with baseline raw P-tau217 concentrations of 0.2, 0.4, 0.6, and 0.8 pg/mL. For modeling, these raw values were transformed using the same log₁₀ and z-standardization procedures. Overlapping curves indicate similar adjusted survival probabilities at a given time point.

**Supplementary Figure 4. Adjusted survival probabilities showing the temporal dynamics of plasma P-tau217 positivity, defined by a sensitivity-maximizing cutoff, in predicting cognitive impairment across cohorts, overall and stratified by *APOE*-ε4 status.**

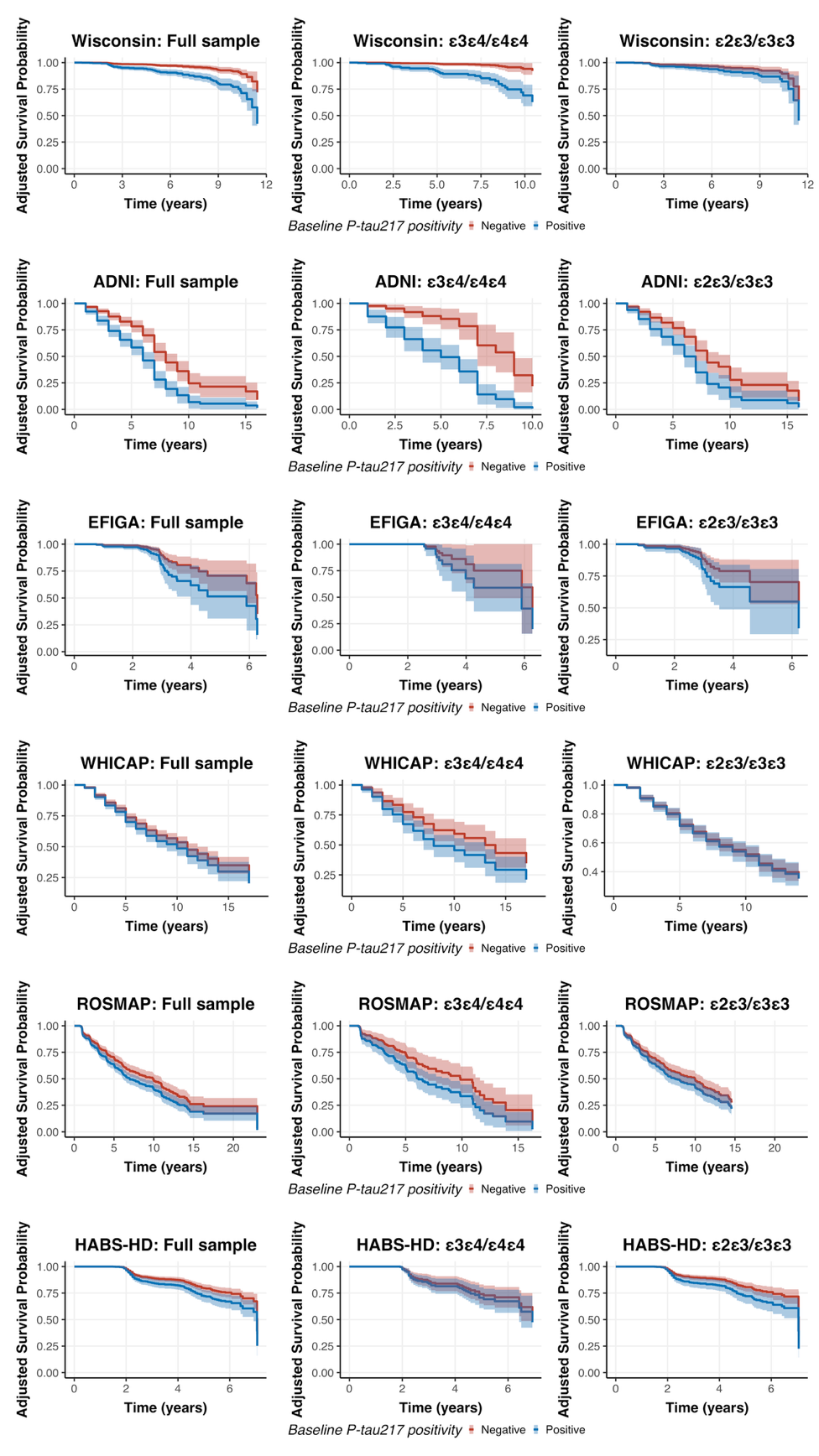

**Supplementary Figure 4** presents cohort-specific model-based curves of adjusted survival probability for cognitive impairment, shown for the full sample and stratified by *APOE*-ε4 status, using baseline P-tau217 positivity as the predictor. P-tau217 positivity was defined using cohort-specific thresholds that maximized sensitivity. Because established cutoffs were not available for EFIGA, ROSMAP, and HABS-HD, we applied the WHICAP cutoff as a proxy, given that these cohorts are similarly multi-ancestry and predominantly non-White populations. The adjusted survival probability curves represent the estimated probability of remaining cognitively unimpaired over time after covariate adjustment and were derived from Cox proportional hazards models with incident cognitive impairment (i.e., MCI or AD) as the outcome. All models were adjusted for age, sex, education, ethnicity, and *APOE* genotype. Stratification was applied in models where the proportional hazards assumption was violated. Within each cohort, the x-axis represents follow-up time (years), and the y-axis shows the adjusted survival probability, that is, the estimated probability of remaining cognitively unimpaired at each follow-up time for each P-tau217 group. Overlapping curves indicate similar adjusted survival probabilities between groups at a given time point.

**Supplementary Figure 5. Adjusted survival probabilities showing the temporal dynamics of plasma P-tau217 positivity, defined by a specificity-maximizing cutoff, in predicting cognitive impairment across cohorts, overall and stratified by *APOE*-ε4 status.**

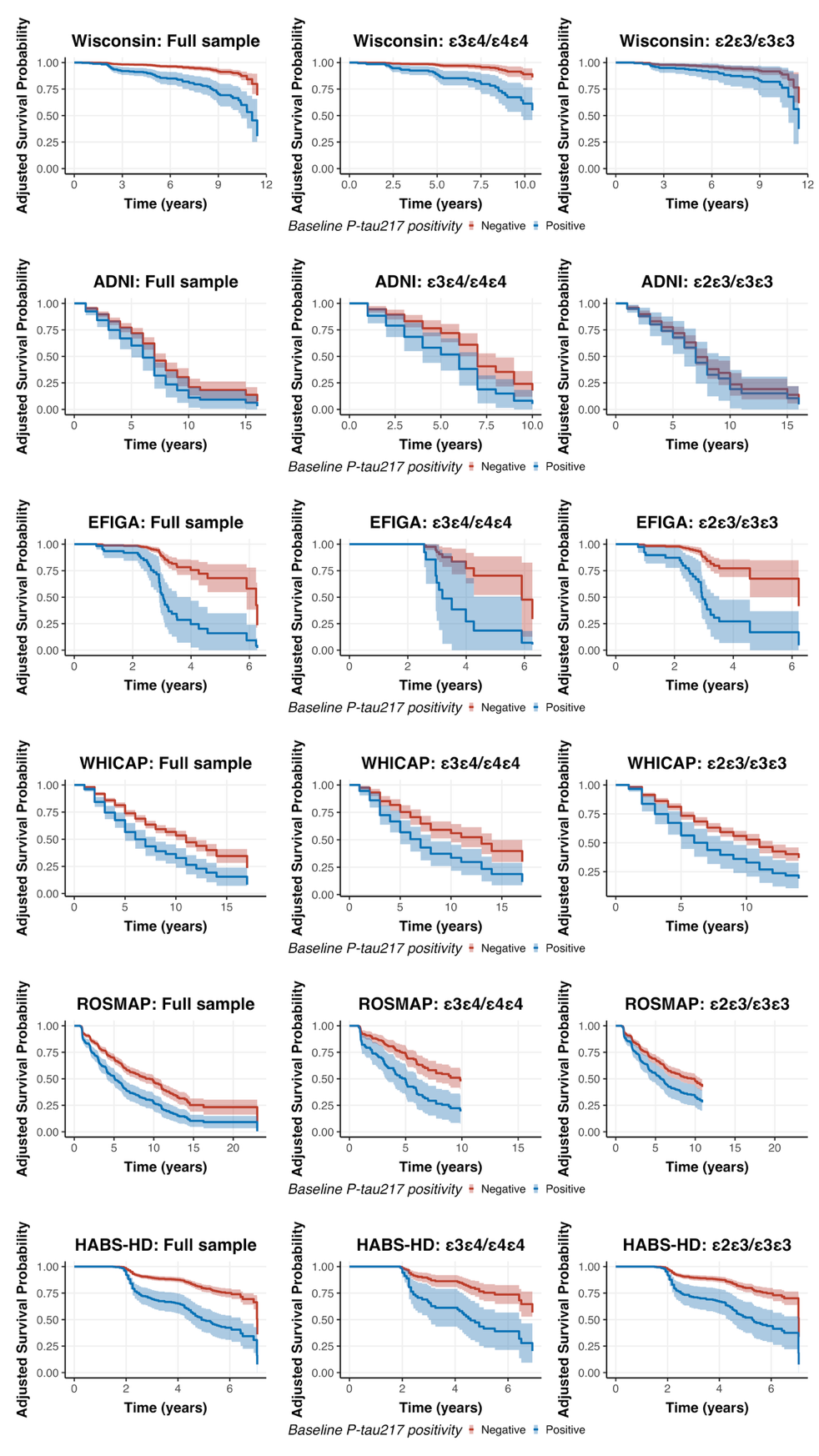

**Supplementary Figure 5** presents cohort-specific model-based curves of adjusted survival probability for cognitive impairment, shown for the full sample and stratified by *APOE*-ε4 status, using baseline P-tau217 positivity as the predictor. P-tau217 positivity was defined using cohort-specific thresholds that maximized specificity. Because established cutoffs were not available for EFIGA, ROSMAP, and HABS-HD, we applied the WHICAP cutoff as a proxy, given that these cohorts are similarly multi-ancestry and predominantly non-White populations. The adjusted survival probability curves represent the estimated probability of remaining cognitively unimpaired over time after covariate adjustment and were derived from Cox proportional hazards models with incident cognitive impairment (i.e., MCI or AD) as the outcome. All models were adjusted for age, sex, education, ethnicity, and *APOE* genotype. Stratification was applied in models where the proportional hazards assumption was violated. Within each cohort, the x-axis represents follow-up time (years), and the y-axis shows the adjusted survival probability, that is, the estimated probability of remaining cognitively unimpaired at each follow-up time for each P-tau217 group. Overlapping curves indicate similar adjusted survival probabilities between groups at a given time point.

**Supplementary Figure 6. Cohort-specific model-based restricted mean survival time (RMST) curves showing time spent cognitively unimpaired by continuous baseline P-tau217 levels.**

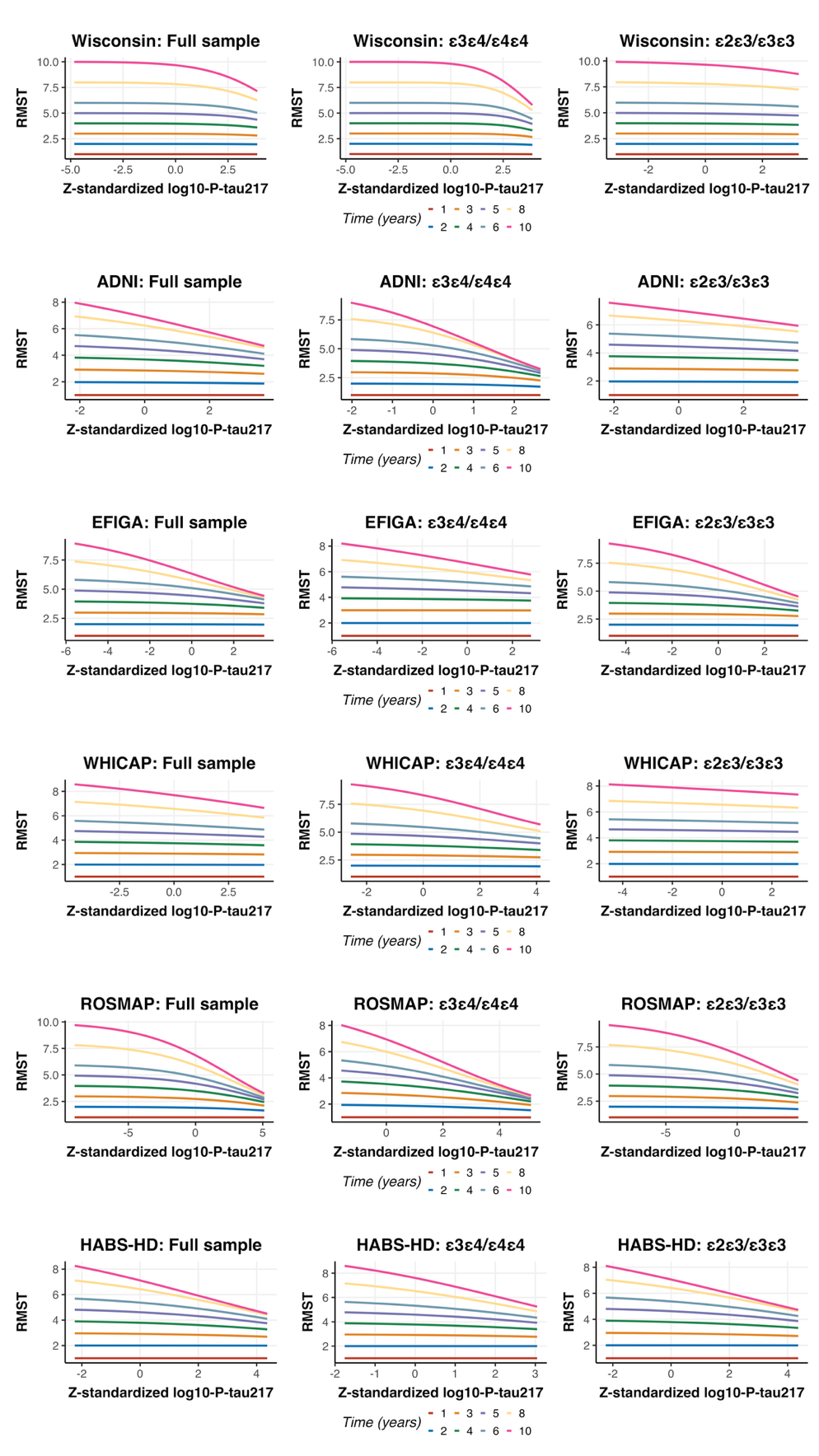

**Supplementary Figure 6** shows cohort-specific model-based restricted mean survival time (RMST) curves for time spent cognitively unimpaired, presented for the full sample and stratified by *APOE*-ε4 status, using baseline continuous P-tau217 levels as predictors. Continuous P-tau217 values were harmonized using log₁₀ transformation followed by z-standardization. RMST curves represent the estimated average duration participants remained cognitively unimpaired and were derived from Cox proportional hazards models with incident cognitive impairment (i.e., MCI or AD) as the outcome. All models were adjusted for age, sex, education, ethnicity, and *APOE* genotype. Stratification was applied in models where the proportional hazards assumption was violated. Within each cohort, the x-axis represents harmonized continuous P‑tau217 levels, and each curve corresponds to a specific follow-up year. The y-axis indicates RMST across baseline P‑tau217 values. Flat (horizontal) curves at early follow-up years reflect minimal differences in RMST between individuals with low versus high P‑tau217. At longer follow-up durations, curves slope downward, indicating shorter cognitively unimpaired periods for those with higher baseline P‑tau217.

**Supplementary Figure 7. Cohort-specific model-based restricted mean survival time (RMST) curves showing time spent cognitively unimpaired by baseline P-tau217 positivity, defined using a cutoff that maximizes sensitivity.**

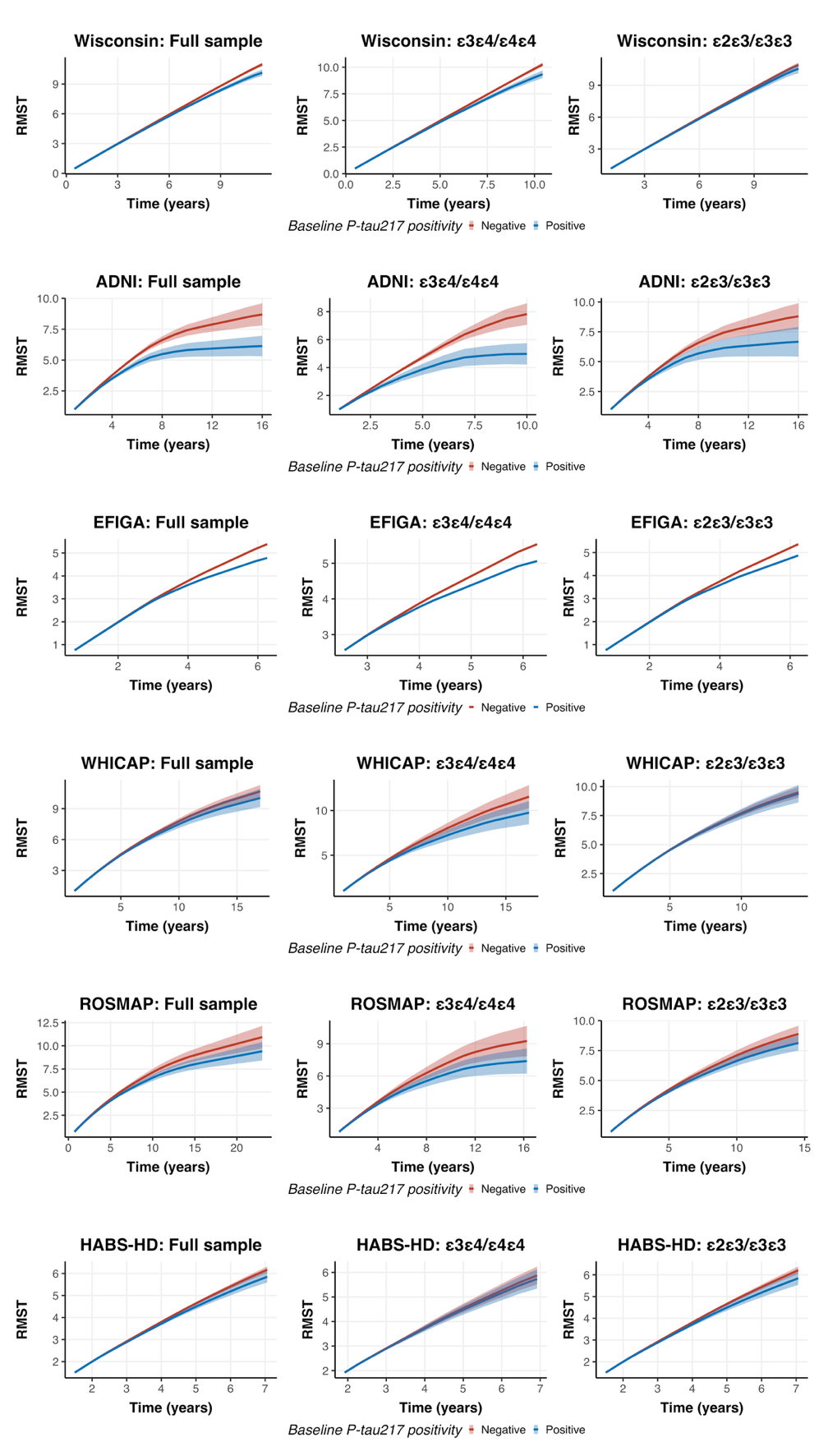

**Supplementary Figure 7** shows cohort-specific model-based restricted mean survival time (RMST) curves for time spent cognitively unimpaired, presented for the full sample and stratified by *APOE*-ε4 status, using baseline P-tau217 positivity as the predictor. P-tau217 positivity was defined using cohort-specific thresholds that maximized sensitivity. Because established cutoffs were not available for EFIGA, ROSMAP, and HABS-HD, we applied the WHICAP cutoff as a proxy, given that these cohorts are similarly multi-ancestry and predominantly non-White populations. RMST curves represent the estimated average duration participants remained cognitively unimpaired and were derived from Cox proportional hazards models with incident cognitive impairment (i.e., MCI or AD) as the outcome. All models were adjusted for age, sex, education, ethnicity, and *APOE* genotype. Within each cohort, the x-axis represents follow-up time (years), and the y-axis shows RMST, defined as the average time participants remained cognitively unimpaired up to each follow-up time point for each P-tau217 group. Overlapping curves indicate similar RMST estimates between groups at a given time point. Confidence intervals were estimated using 1,000 bootstrap resamples. In the EFIGA cohort, limited event/exposure counts and shorter follow-up duration (<10 years) precluded estimation of confidence intervals.

**Supplementary Figure 8. Cohort-specific model-based restricted mean survival time (RMST) curves showing time spent cognitively unimpaired by baseline P-tau217 positivity, defined using a cutoff that maximizes specificity.**

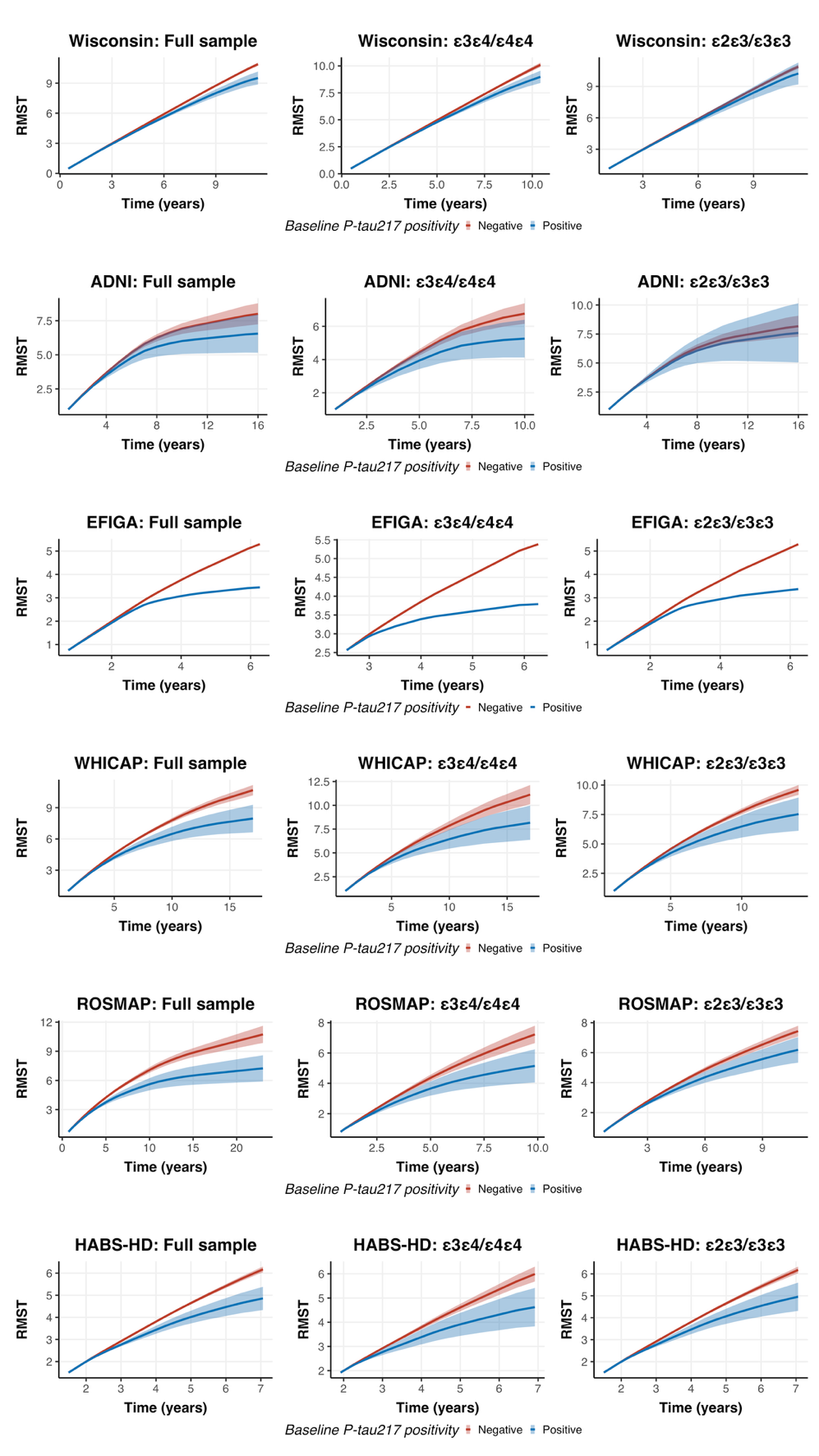

**Supplementary Figure 8** shows cohort-specific model-based restricted mean survival time (RMST) curves for time spent cognitively unimpaired, presented for the full sample and stratified by *APOE*-ε4 status, using baseline P-tau217 positivity as the predictor. P-tau217 positivity was defined using cohort-specific thresholds that maximized specificity. Because established cutoffs were not available for EFIGA, ROSMAP, and HABS-HD, we applied the WHICAP cutoff as a proxy, given that these cohorts are similarly multi-ancestry and predominantly non-White populations. RMST curves represent the estimated average duration participants remained cognitively unimpaired and were derived from Cox proportional hazards models with incident cognitive impairment (i.e., MCI or AD) as the outcome. All models were adjusted for age, sex, education, ethnicity, and *APOE* genotype. Within each cohort, the x-axis represents follow-up time (years), and the y-axis shows RMST, defined as the average time participants remained cognitively unimpaired up to each follow-up time point for each P-tau217 group. Overlapping curves indicate similar RMST estimates between groups at a given time point. Confidence intervals were estimated using 1,000 bootstrap resamples. In the EFIGA cohort, limited event/exposure counts and shorter follow-up duration (<10 years) precluded estimation of confidence intervals.

**Supplementary Figure 9. Baseline and longitudinal prognostic performance of *APOE*-ε4 carrier status and plasma P-tau217 positivity, including P-tau217 stratified by *APOE*-ε4 carrier status, for cognitive impairment in the pooled cohort, as assessed by AUC, incremental R², and Harrell’s C-index.**

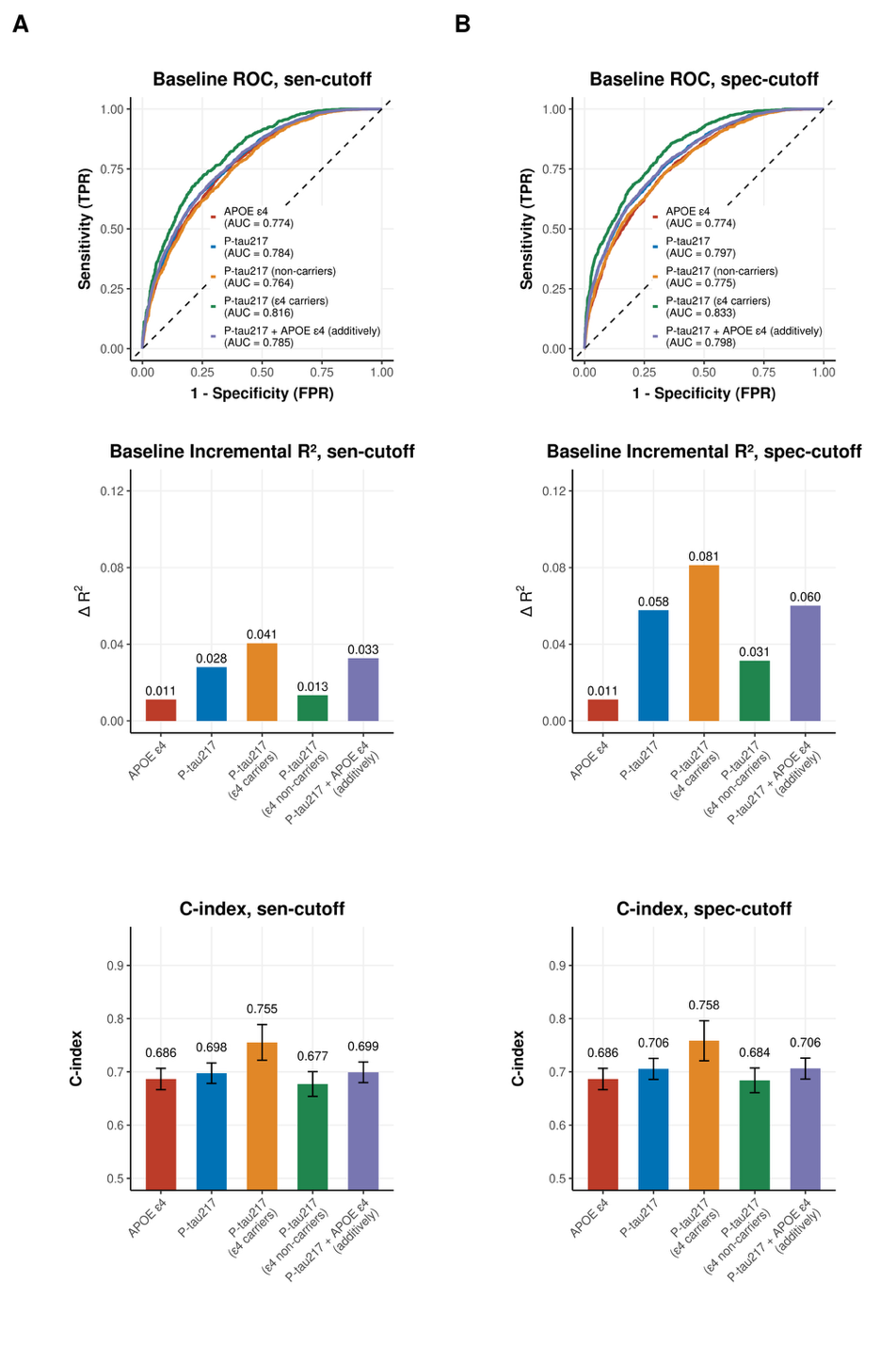

**Supplementary Figure 9** shows baseline and longitudinal prognostic performance of plasma P-tau217 positivity for cognitive impairment in the pooled cohort. Receiver operating characteristic (ROC) curves were obtained from logistic regression models adjusted for age, sex, education, cohort, and ethnic group. Models included *APOE*-ε4 status with covariates; plasma P-tau217 with covariates; plasma P-tau217 and *APOE*-ε4 entered additively with covariates; and the plasma P-tau217 with covariates fitted separately within *APOE*-ε4 carrier and non-carrier groups. Incremental Nagelkerke’s R² was defined as the increase in model fit when adding the predictor or the predictors of interest to the covariate-only model. Harrell’s C-indices were obtained from Cox proportional hazards models adjusted for the same covariates and specified analogously. P‑tau217 positivity was defined using cohort-specific thresholds that maximized sensitivity (Supplementary Figure 9A) or specificity (Supplementary Figure 9B). Because established cutoffs were not available for EFIGA, ROSMAP, and HABS-HD, we applied the WHICAP cutoff as a proxy, given that these cohorts are similarly multi-ancestry and predominantly non-White populations.

**Supplementary Figure 10. Prognostic performance of plasma P-tau217 for cognitive impairment at baseline and longitudinally, assessed using AUC, incremental R², and C-index. Performance is shown when P-tau217 is modeled as a continuous measure (A) or as a binary positivity variable using cutoffs that maximize sensitivity (B) or specificity (C) in the Wisconsin dataset.**

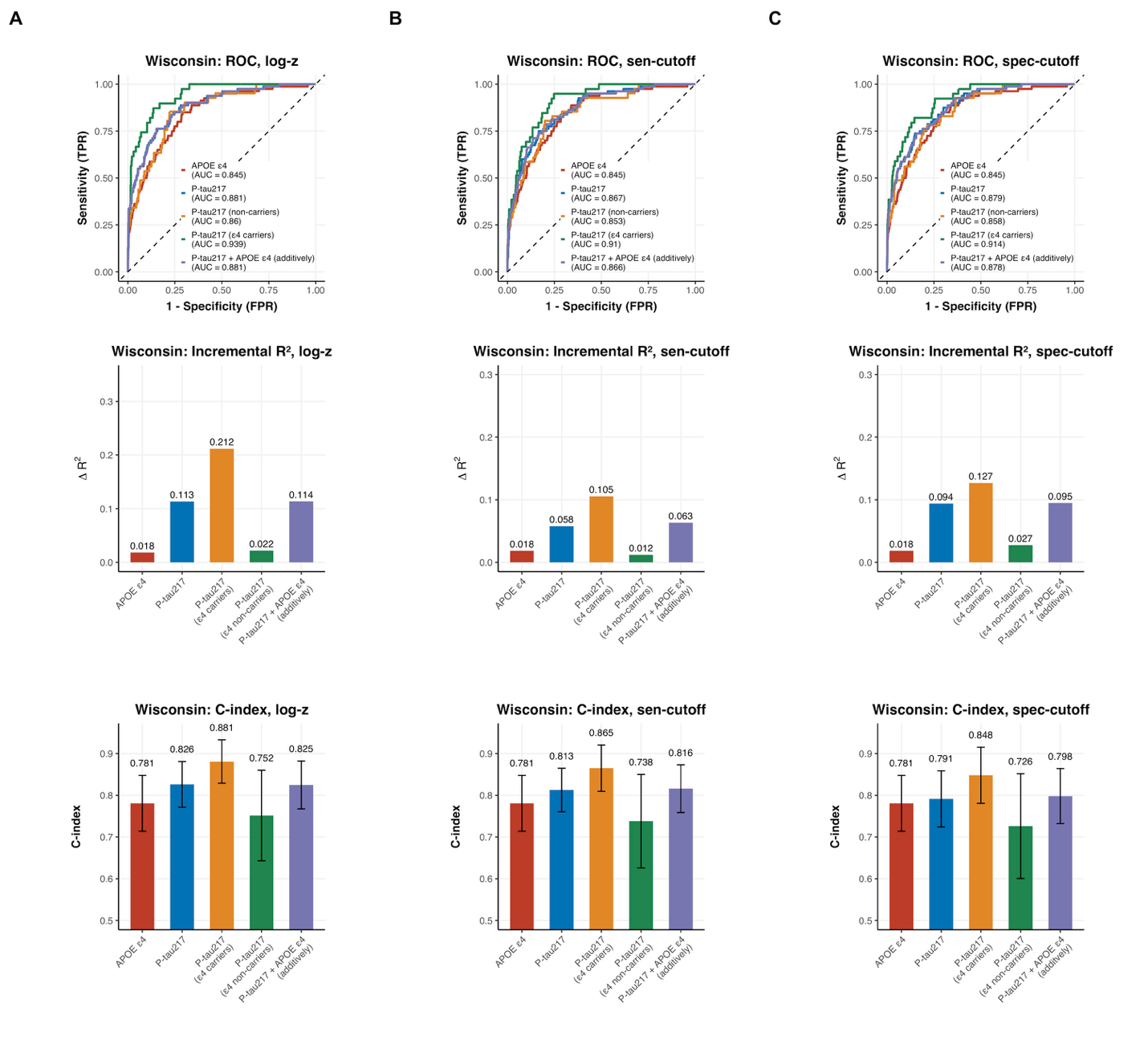

**Supplementary Figure 10** showed baseline and longitudinal prognostic performance of plasma P-tau217 levels and positivity for cognitive impairment in the Wisconsin cohort. Supplementary Figure 10A shows results when P-tau217 is modeled as a continuous variable, supplementary Figure 10B shows results when P-tau217 positivity is defined using the sensitivity-based cut-off, and supplementary Figure 10C presents results when positivity is defined using the specificity-based cut-off. Receiver operating characteristic (ROC) curves were obtained from logistic regression models adjusted for age, sex, education, and ethnic group. The *APOE*-ε4 model included *APOE*-ε4 status with covariates; the P-tau217 model included plasma P-tau217 with covariates; and the P-tau217 (ε4 carriers) and P-tau217 (non-carriers) models included plasma P-tau217 with covariates, fitted separately within *APOE*-ε4 carrier and non-carrier groups. Incremental Nagelkerke’s R² was defined as the increase in model fit when adding the predictor of interest to the covariate-only model. Harrell’s C-indices were obtained from Cox proportional hazards models adjusted for the same covariates. The *APOE*-ε4 model included *APOE-*ε4 status with covariates; the P-tau217 model included plasma P-tau217 with covariates; and the P-tau217 (ε4 carriers) and P-tau217 (non-carriers) models included plasma P-tau217 with covariates, fitted separately within *APOE*-ε4 carrier and non-carrier groups.

**Supplementary Figure 11. Prognostic performance of plasma P-tau217 for cognitive impairment at baseline and longitudinally, assessed using AUC, incremental R², and C-index. Performance is shown when P-tau217 is modeled as a continuous measure (A) or as a binary positivity variable using cutoffs that maximize sensitivity (B) or specificity (C) in the ADNI dataset.**

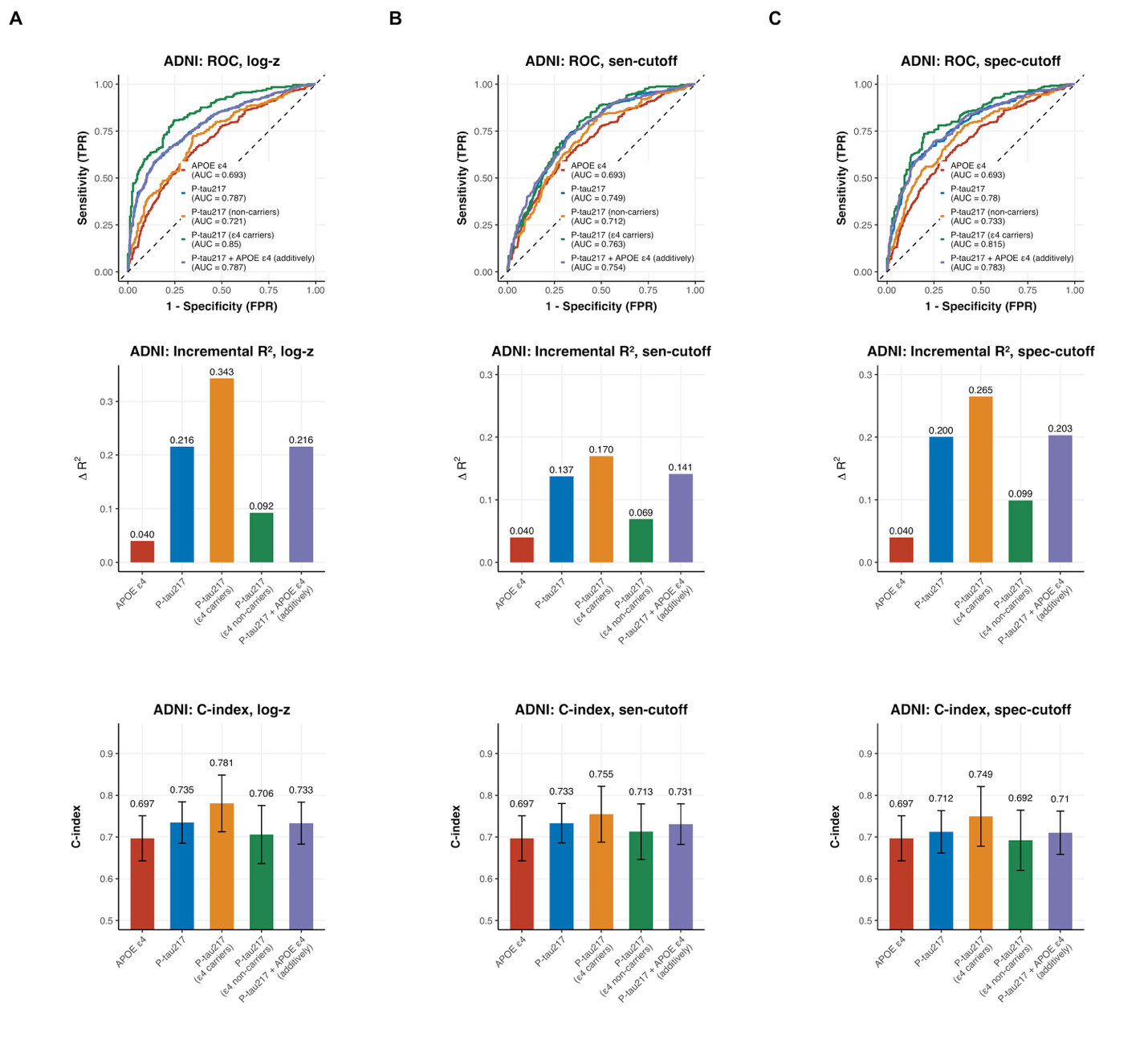

**Supplementary Figure 11** showed baseline and longitudinal prognostic performance of plasma P-tau217 levels and positivity for cognitive impairment in the ADNI cohort. Supplementary Figure 11A shows results when P-tau217 is modeled as a continuous variable, supplementary Figure 11B shows results when P-tau217 positivity is defined using the sensitivity-based cut-off, and supplementary Figure 11C presents results when positivity is defined using the specificity-based cut-off. Receiver operating characteristic (ROC) curves were obtained from logistic regression models adjusted for age, sex, education, and ethnic group. The *APOE*-ε4 model included *APOE*-ε4 status with covariates; the P-tau217 model included plasma P-tau217 with covariates; and the P-tau217 (ε4 carriers) and P-tau217 (non-carriers) models included plasma P-tau217 with covariates, fitted separately within *APOE*-ε4 carrier and non-carrier groups. Incremental Nagelkerke’s R² was defined as the increase in model fit when adding the predictor of interest to the covariate-only model. Harrell’s C-indices were obtained from Cox proportional hazards models adjusted for the same covariates. The *APOE*-ε4 model included *APOE-*ε4 status with covariates; the P-tau217 model included plasma P-tau217 with covariates; and the P-tau217 (ε4 carriers) and P-tau217 (non-carriers) models included plasma P-tau217 with covariates, fitted separately within *APOE*-ε4 carrier and non-carrier groups.

**Supplementary Figure 12. Prognostic performance of plasma P-tau217 for cognitive impairment at baseline and longitudinally, assessed using AUC, incremental R², and C-index. Performance is shown when P-tau217 is modeled as a continuous measure (A) or as a binary positivity variable using cutoffs that maximize sensitivity (B) or specificity (C) in the EFIGA dataset.**

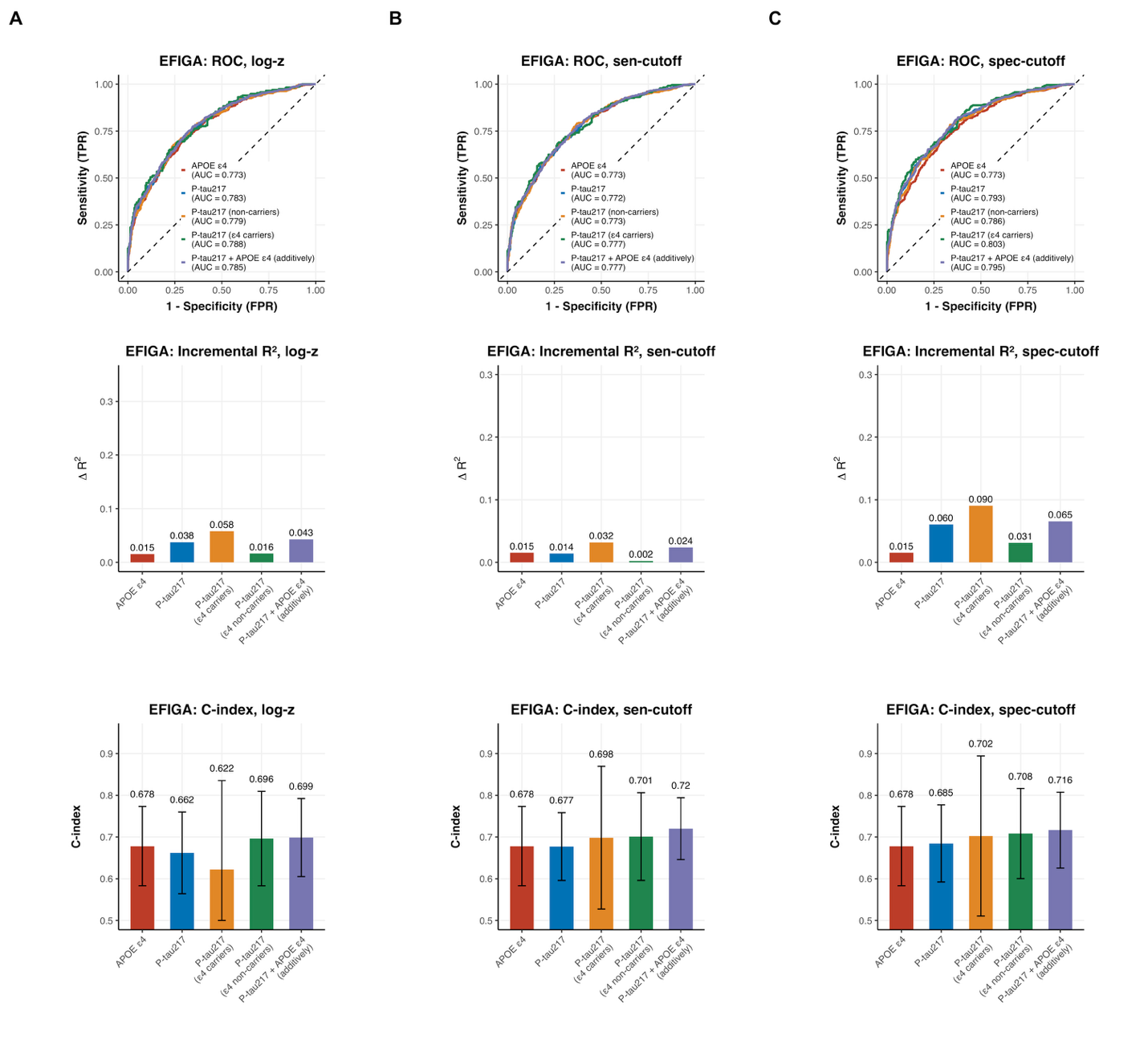

**Supplementary Figure 12** showed baseline and longitudinal prognostic performance of plasma P-tau217 levels and positivity for cognitive impairment in the EFIGA cohort. Because established cutoffs were not available for EFIGA, ROSMAP, and HABS-HD, we applied the WHICAP cutoff as a proxy, given that these cohorts are similarly multi-ancestry and predominantly non-White populations. Supplementary Figure 12A shows results when P-tau217 is modeled as a continuous variable, supplementary Figure 12B shows results when P-tau217 positivity is defined using the sensitivity-based cut-off, and supplementary Figure 12C presents results when positivity is defined using the specificity-based cut-off. Receiver operating characteristic (ROC) curves were obtained from logistic regression models adjusted for age, sex, and education. The *APOE*-ε4 model included *APOE*-ε4 status with covariates; the P-tau217 model included plasma P-tau217 with covariates; and the P-tau217 (ε4 carriers) and P-tau217 (non-carriers) models included plasma P-tau217 with covariates, fitted separately within *APOE*-ε4 carrier and non-carrier groups. Incremental Nagelkerke’s R² was defined as the increase in model fit when adding the predictor of interest to the covariate-only model. Harrell’s C-indices were obtained from Cox proportional hazards models adjusted for the same covariates. The *APOE*-ε4 model included *APOE-*ε4 status with covariates; the P-tau217 model included plasma P-tau217 with covariates; and the P-tau217 (ε4 carriers) and P-tau217 (non-carriers) models included plasma P-tau217 with covariates, fitted separately within *APOE*-ε4 carrier and non-carrier groups.

**Supplementary Figure 13. Prognostic performance of plasma P-tau217 for cognitive impairment at baseline and longitudinally, assessed using AUC, incremental R², and C-index. Performance is shown when P-tau217 is modeled as a continuous measure (A) or as a binary positivity variable using cutoffs that maximize sensitivity (B) or specificity (C) in the WHICAP dataset.**

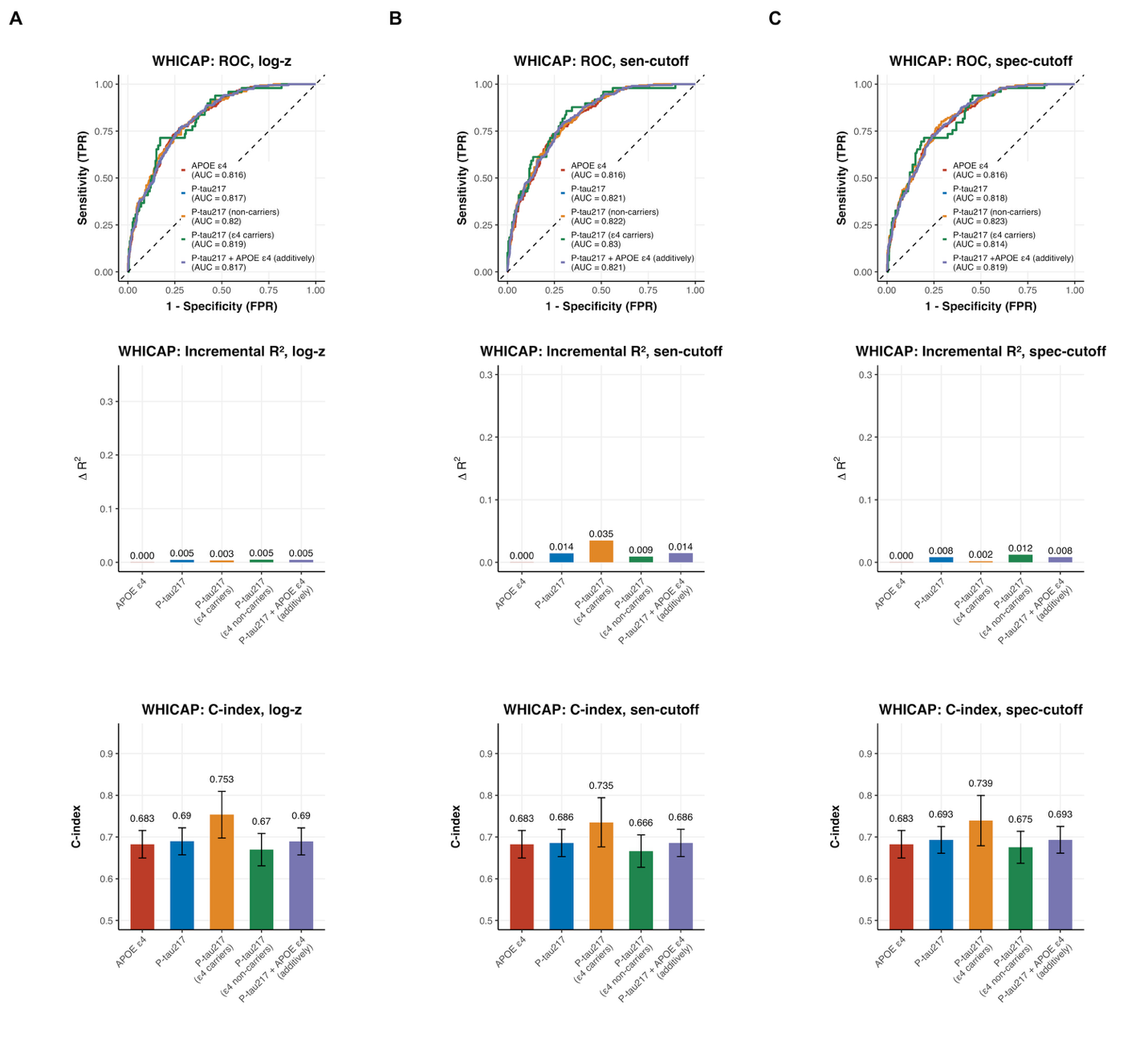

**Supplementary Figure 13** showed baseline and longitudinal prognostic performance of plasma P-tau217 levels and positivity for cognitive impairment in the WHICAP cohort. Supplementary Figure 13A shows results when P-tau217 is modeled as a continuous variable, supplementary Figure 13B shows results when P-tau217 positivity is defined using the sensitivity-based cut-off, and supplementary Figure 13C presents results when positivity is defined using the specificity-based cut-off. Receiver operating characteristic (ROC) curves were obtained from logistic regression models adjusted for age, sex, education, and ethnic group. The *APOE*-ε4 model included *APOE*-ε4 status with covariates; the P-tau217 model included plasma P-tau217 with covariates; and the P-tau217 (ε4 carriers) and P-tau217 (non-carriers) models included plasma P-tau217 with covariates, fitted separately within *APOE*-ε4 carrier and non-carrier groups. Incremental Nagelkerke’s R² was defined as the increase in model fit when adding the predictor of interest to the covariate-only model. Harrell’s C-indices were obtained from Cox proportional hazards models adjusted for the same covariates. The *APOE*-ε4 model included *APOE-*ε4 status with covariates; the P-tau217 model included plasma P-tau217 with covariates; and the P-tau217 (ε4 carriers) and P-tau217 (non-carriers) models included plasma P-tau217 with covariates, fitted separately within *APOE*-ε4 carrier and non-carrier groups.

**Supplementary Figure 14. Prognostic performance of plasma P-tau217 for cognitive impairment at baseline and longitudinally, assessed using AUC, incremental R², and C-index. Performance is shown when P-tau217 is modeled as a continuous measure (A) or as a binary positivity variable using cutoffs that maximize sensitivity (B) or specificity (C) in the ROSMAP dataset.**

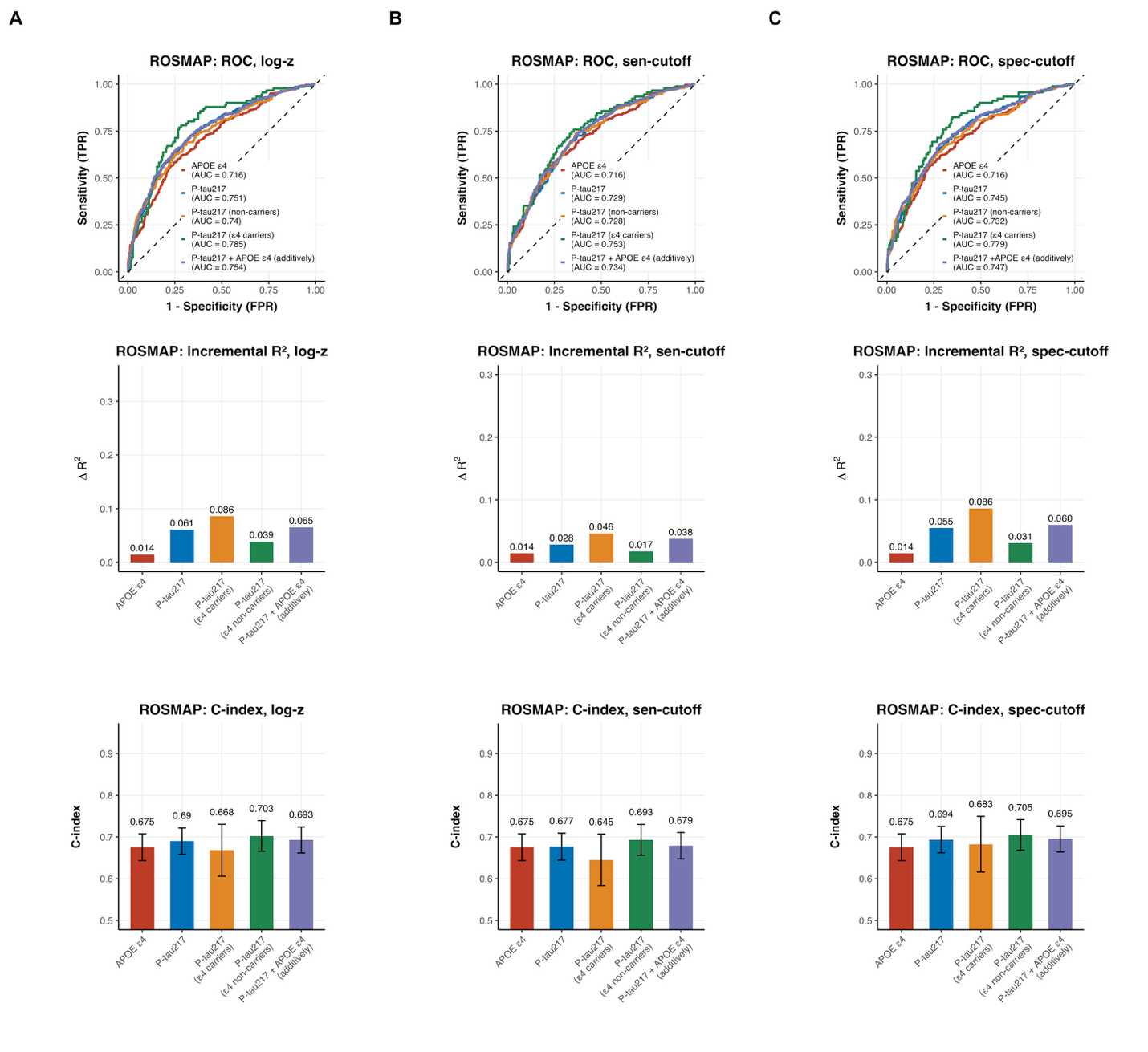

**Supplementary Figure 14** showed baseline and longitudinal prognostic performance of plasma P-tau217 levels and positivity for cognitive impairment in the ROSMAP cohort. Because established cutoffs were not available for EFIGA, ROSMAP, and HABS-HD, we applied the WHICAP cutoff as a proxy, given that these cohorts are similarly multi-ancestry and predominantly non-White populations. Supplementary Figure 14A shows results when P-tau217 is modeled as a continuous variable, supplementary Figure 14B shows results when P-tau217 positivity is defined using the sensitivity-based cut-off, and supplementary Figure 14C presents results when positivity is defined using the specificity-based cut-off. Receiver operating characteristic (ROC) curves were obtained from logistic regression models adjusted for age, sex, education, and ethnic group. The *APOE*-ε4 model included *APOE*-ε4 status with covariates; the P-tau217 model included plasma P-tau217 with covariates; and the P-tau217 (ε4 carriers) and P-tau217 (non-carriers) models included plasma P-tau217 with covariates, fitted separately within *APOE*-ε4 carrier and non-carrier groups. Incremental Nagelkerke’s R² was defined as the increase in model fit when adding the predictor of interest to the covariate-only model. Harrell’s C-indices were obtained from Cox proportional hazards models adjusted for the same covariates. The *APOE*-ε4 model included *APOE-*ε4 status with covariates; the P-tau217 model included plasma P-tau217 with covariates; and the P-tau217 (ε4 carriers) and P-tau217 (non-carriers) models included plasma P-tau217 with covariates, fitted separately within *APOE*-ε4 carrier and non-carrier groups.

**Supplementary Figure 15. Prognostic performance of plasma P-tau217 for cognitive impairment at baseline and longitudinally, assessed using AUC, incremental R², and C-index. Performance is shown when P-tau217 is modeled as a continuous measure (A) or as a binary positivity variable using cutoffs that maximize sensitivity (B) or specificity (C) in the HABS-HD dataset.**

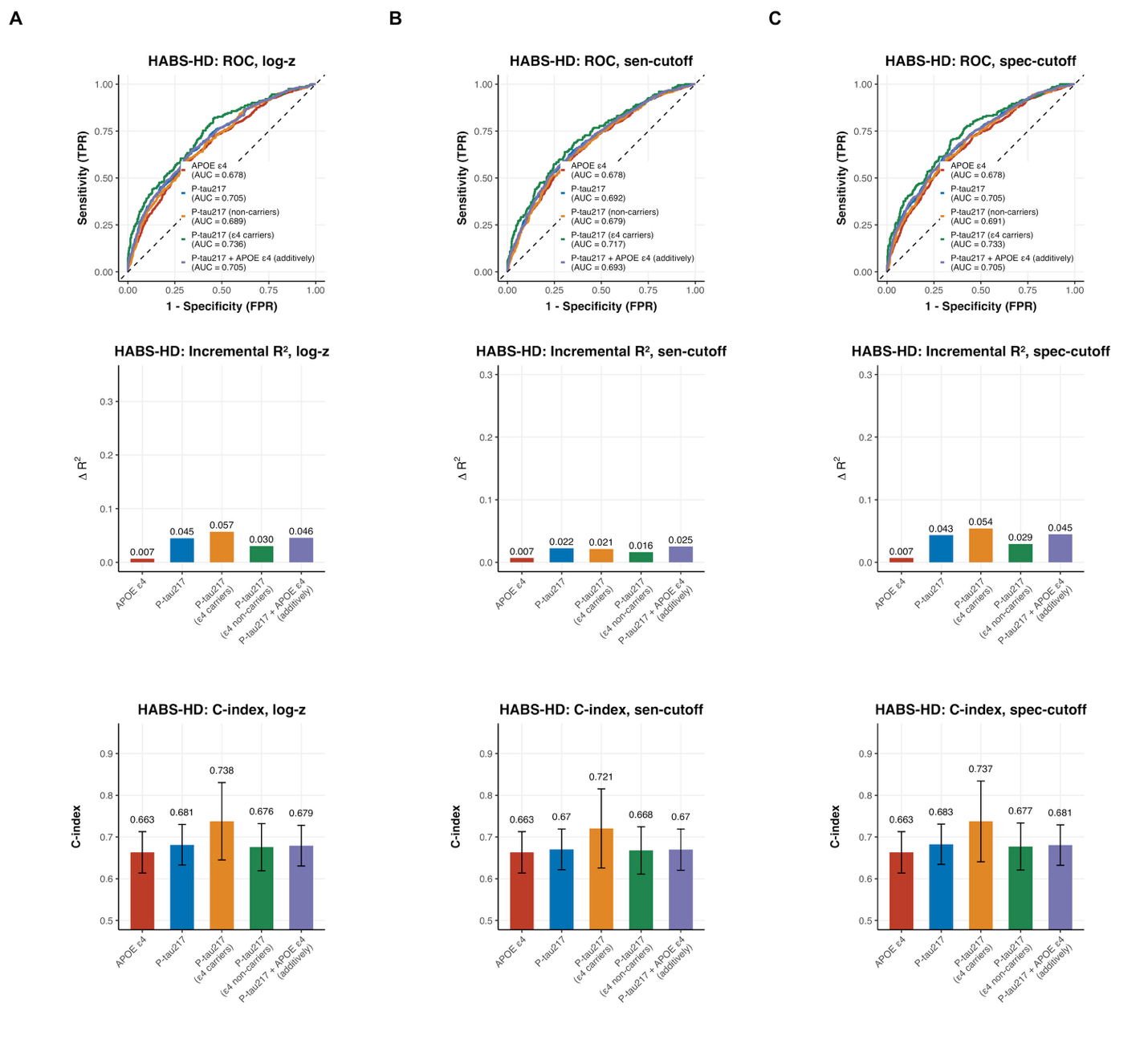

**Supplementary Figure 15** showed baseline and longitudinal prognostic performance of plasma P-tau217 levels and positivity for cognitive impairment in the HABS-HD cohort. Because established cutoffs were not available for EFIGA, ROSMAP, and HABS-HD, we applied the WHICAP cutoff as a proxy, given that these cohorts are similarly multi-ancestry and predominantly non-White populations. Supplementary Figure 15A shows results when P-tau217 is modeled as a continuous variable, supplementary Figure 15B shows results when P-tau217 positivity is defined using the sensitivity-based cut-off, and supplementary Figure 15C presents results when positivity is defined using the specificity-based cut-off. Receiver operating characteristic (ROC) curves were obtained from logistic regression models adjusted for age, sex, education, and ethnic group. The *APOE*-ε4 model included *APOE*-ε4 status with covariates; the P-tau217 model included plasma P-tau217 with covariates; and the P-tau217 (ε4 carriers) and P-tau217 (non-carriers) models included plasma P-tau217 with covariates, fitted separately within *APOE*-ε4 carrier and non-carrier groups. Incremental Nagelkerke’s R² was defined as the increase in model fit when adding the predictor of interest to the covariate-only model. Harrell’s C-indices were obtained from Cox proportional hazards models adjusted for the same covariates. The *APOE*-ε4 model included *APOE-*ε4 status with covariates; the P-tau217 model included plasma P-tau217 with covariates; and the P-tau217 (ε4 carriers) and P-tau217 (non-carriers) models included plasma P-tau217 with covariates, fitted separately within *APOE*-ε4 carrier and non-carrier groups.

**Supplementary Figure 16. Adjusted survival probabilities illustrating the temporal dynamics of plasma P-tau217 positivity, defined using a Wisconsin-derived cutoff that maximized sensitivity (A) and specificity (B) for predicting incident cognitive impairment in the pooled cohort, overall and stratified by *APOE*-ε4 status, together with model-based restricted mean survival time (RMST) curves showing time spent cognitively unimpaired stratified by baseline P-tau217 positivity.**

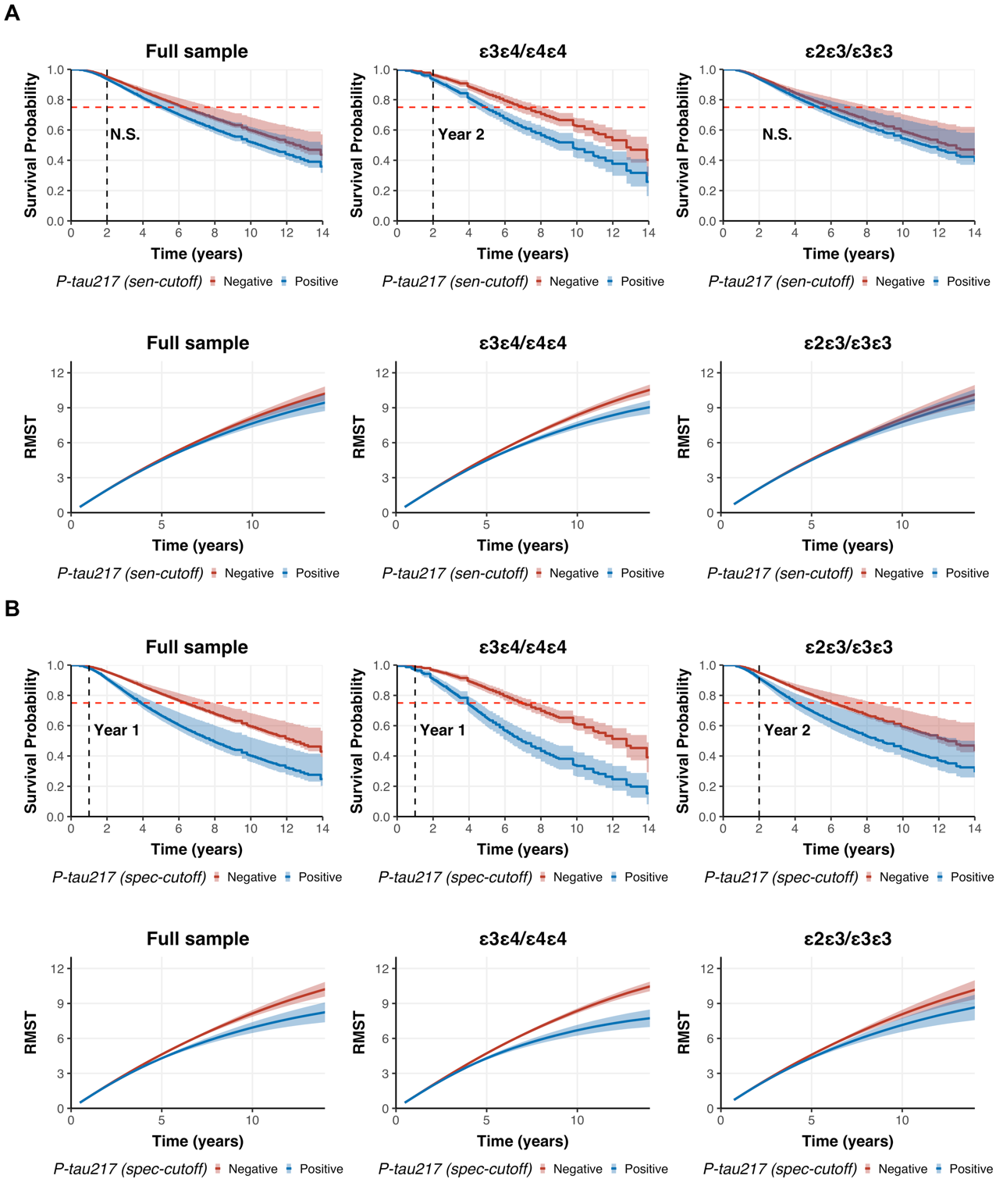

**Supplementary Figure 16** presents adjusted survival probabilities illustrating the temporal dynamics of plasma P-tau217 positivity, defined using Wisconsin-derived cutoffs that maximized sensitivity (A) and specificity (B) for predicting incident cognitive impairment in the pooled cohort, overall and stratified by *APOE*-ε4 status, together with model-based restricted mean survival time (RMST) curves showing time spent cognitively unimpaired stratified by baseline P-tau217 positivity. To maximize statistical power for the adjusted survival analyses, data from all cohorts were combined. P-tau217 positivity was defined within each cohort using thresholds that maximized sensitivity or specificity, as determined in the Wisconsin cohort, and then pooled across cohorts. Because the proportional hazards assumption was violated in the combined sample, a pseudo-value approach (implemented using the adjustedCurves package in R) was applied when modeling P-tau217 as a binary positivity measure. This method estimates survival probabilities at prespecified time points, adjusts for covariates (age, sex, education, cohort, ethnicity, and *APOE* genotype), and averages predictions across groups. Confidence intervals were obtained using 1,000 bootstrap resamples.

We also present model-based RMST curves for time spent cognitively unimpaired, shown for the full sample and stratified by *APOE*-ε4 status, using binary P-tau217 positivity thresholds that maximized sensitivity or specificity as predictors, as determined from the Wisconsin cohort. RMST curves represent the estimated average duration participants remained cognitively unimpaired and were derived from Cox proportional hazards models with incident cognitive impairment (i.e., MCI or AD) as the outcome. All models were adjusted for age, sex, education, cohort, ethnicity, and *APOE* genotype, with stratification applied when the proportional hazards assumption was violated. For binary P-tau217 measures, the x-axis represents follow-up time (years), and the y-axis shows RMST, defined as the average time participants remained cognitively unimpaired up to each follow-up time point for each P-tau217 group. Overlapping curves indicate similar RMST estimates between groups at a given time point. Confidence intervals were estimated using 1,000 bootstrap resamples.

**Supplementary Figure 17. Baseline and longitudinal prognostic performance of plasma P-tau217 positivity for cognitive impairment in the pooled cohort, as assessed by AUC, incremental R², and Harrell’s C-index, with P-tau217 positivity defined using the Wisconsin cohort cutoff.**

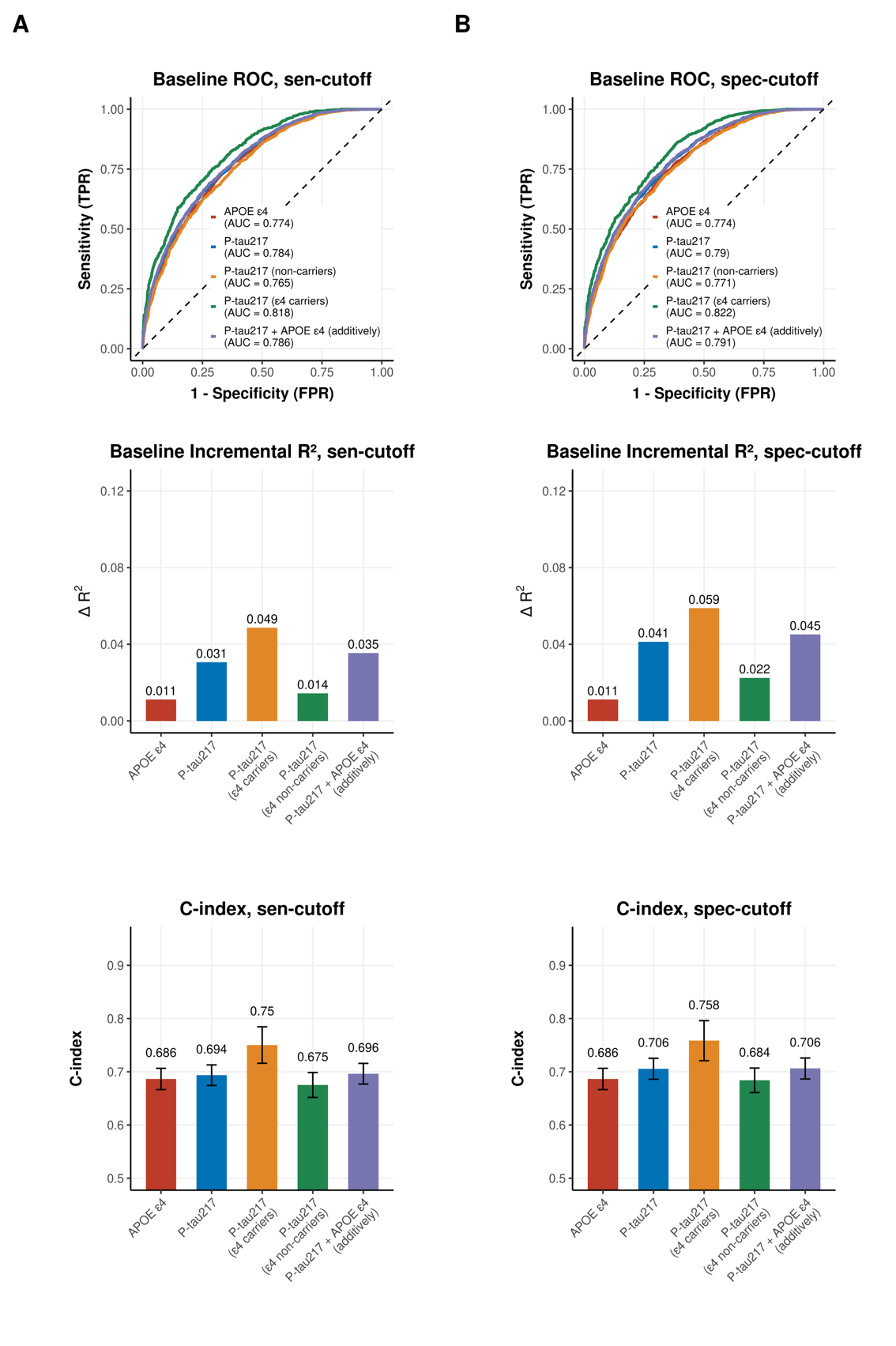

**Supplementary Figure 17** shows the baseline and longitudinal prognostic performance of plasma P-tau217 positivity for cognitive impairment in the pooled cohort, when the cutoff was determined using the Wisconsin cohort. The P-tau217 positivity cutoff was set at 0.63 to maximize specificity and 0.40 to maximize sensitivity, and the same cutoffs were applied uniformly across all cohorts. Supplementary Figure 17A shows results when P-tau217 positivity is defined using the sensitivity-based cut-off, and Supplementary Figure 17B presents results when positivity is defined using the specificity-based cut-off. Receiver operating characteristic (ROC) curves were obtained from logistic regression models adjusted for age, sex, education, cohort, and ethnic group. The *APOE*-ε4 model included *APOE*-ε4 status with covariates; the P-tau217 model included plasma P-tau217 with covariates; and the P-tau217 (ε4 carriers) and P-tau217 (non-carriers) models included plasma P-tau217 with covariates, fitted separately within *APOE*-ε4 carrier and non-carrier groups. Incremental Nagelkerke’s R² was defined as the increase in model fit when adding the predictor of interest to the covariate-only model. Harrell’s C-indices were obtained from Cox proportional hazards models adjusted for the same covariates. The *APOE*-ε4 model included *APOE-*ε4 status with covariates; the P-tau217 model included plasma P-tau217 with covariates; and the P-tau217 (ε4 carriers) and P-tau217 (non-carriers) models included plasma P-tau217 with covariates, fitted separately within *APOE*-ε4 carrier and non-carrier groups.

**Supplementary Figure 18. Predictive performance over follow-up using nonparametric random forest models based on baseline variables with leave-one-cohort-out validation.**

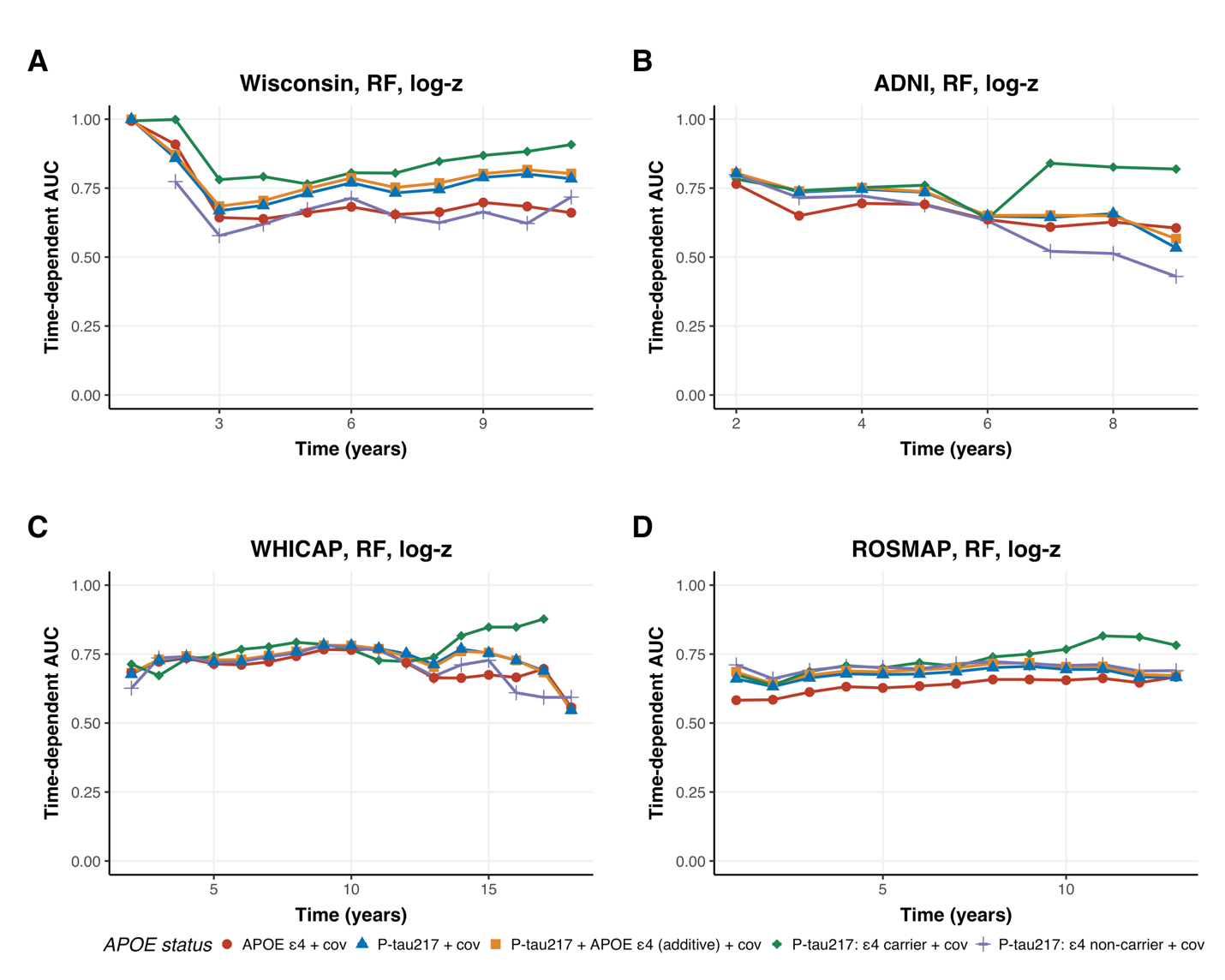

**Supplementary Figure 18** shows predictive performance over follow-up using nonparametric random forest models based on baseline variables with leave-one-cohort-out validation. Continuous P-tau217 was used because continuous predictors are better suited for random forest models than dichotomized variables. Models were trained by combining all cohorts while leaving one cohort out for external validation. We evaluated several models, including *APOE*-ε4 with covariates, P-tau217 with covariates, P-tau217 plus *APOE*-ε4 with covariates, and P-tau217 models fitted separately among ε4 carriers and non-carriers with covariates. Training was performed using all but one cohort, and the held-out cohort was used for testing. All models included age, sex, education, ethnicity, and cohort as covariates in the training data. In the testing cohort, time-dependent AUCs for prediction of cognitive impairment were calculated while adjusting for age, sex, education, and ethnicity. HABS-HD and EFIGA were excluded from testing but included in training because of limited follow-up time.

**Supplementary Figure 19. Cohort-specific and meta-analytic estimates of the association between *APOE*-ε4 carrier status and plasma P‑tau217 levels in the full sample, among cognitively normal and P‑tau217–negative individuals (A), and for *APOE*-ε4 associations with P-tau217 positivity (B)**

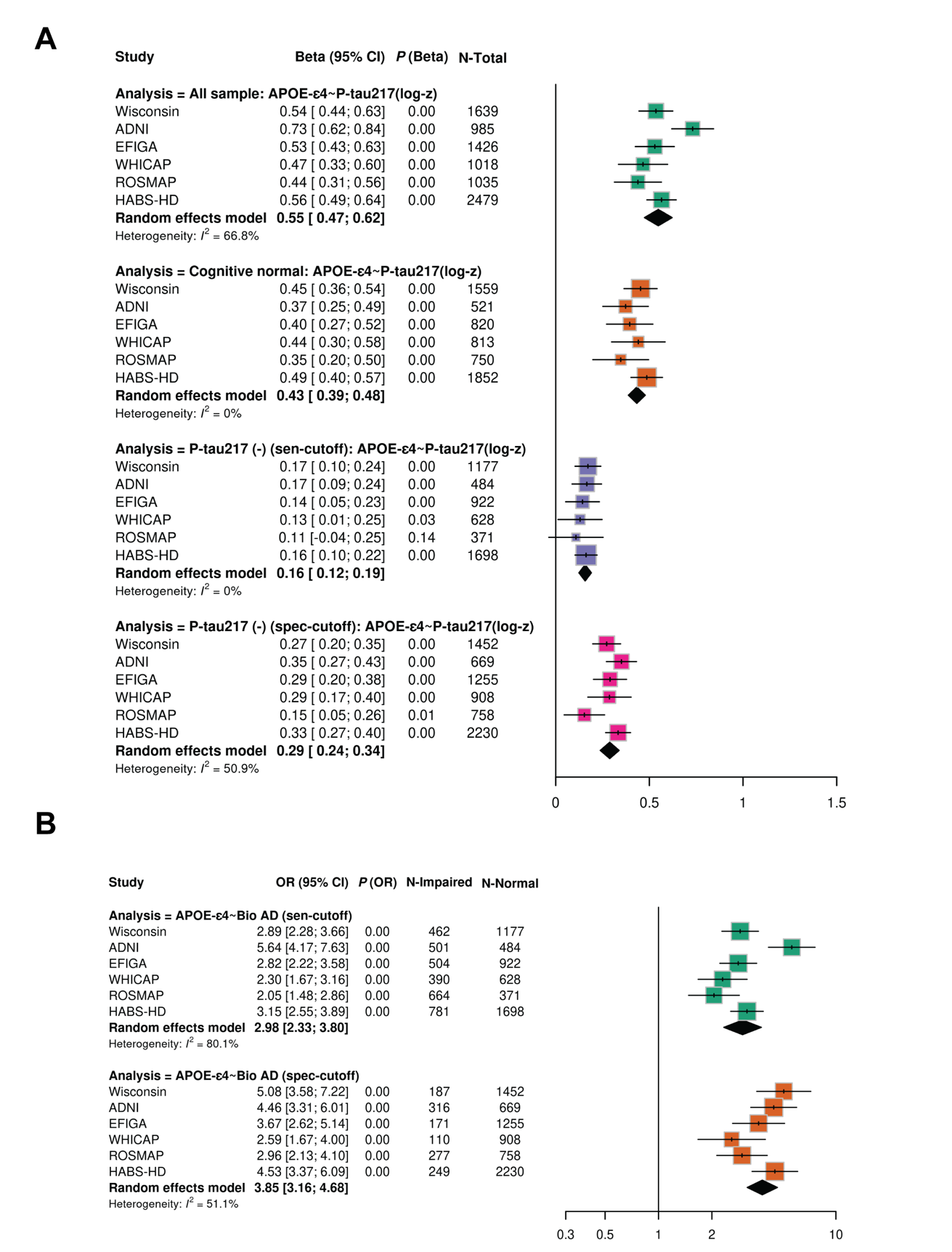

**Supplementary Figure 19** presents cohort-specific and meta-analytic estimates of the association between *APOE*-ε4 carrier status and plasma P‑tau217 levels in the full sample, among cognitively normal and P‑tau217–negative individuals (A), as well as the associations between *APOE*-ε4 and P-tau217 positivity (B). Analyses were conducted at the time of first available biomarker measurement. For cohorts with only cross-sectional data, this corresponds to a single time point; for longitudinal cohorts, it reflects the earliest available biomarker. Plasma P‑tau217 was harmonized using log₁₀ transformation and z-standardization within each cohort. All analyses used linear regression models adjusted for age, sex, education, and ethnicity; Random-effects meta-analyses were conducted using cohort-specific beta estimates. Cognitively normal was defined as the absence of a diagnosis of MCI or AD at the time of biomarker collection. P‑tau217–negative status was defined as having P‑tau217 levels below the cohort-specific threshold that maximized sensitivity or specificity.
